# The effect of lifestyle interventions using behavior change techniques to improve physical activity, sedentary behavior and/or sleep in adults with type 2 diabetes mellitus: a systematic review and meta-analysis of randomized controlled trials

**DOI:** 10.64898/2026.03.24.26349179

**Authors:** Marieke De Craemer, Manon Kinaupenne, Marga Decraene, Lotte Bogaert, Iris Willems

## Abstract

**Introduction/Aim:** Type 2 diabetes (T2D) is a growing global health burden, with lifestyle behaviors playing a key role in its management. Physical activity (PA), sedentary behavior (SB), and sleep are increasingly conceptualized as interdependent components of 24-hour movement behaviors. While behavior change techniques (BCTs) are commonly used to target individual behaviors, their effectiveness across multiple behaviors in adults with T2D remains unclear. This systematic review and meta-analysis aimed to evaluate the effectiveness of behavior change interventions incorporating BCTs on PA, SB, and sleep outcomes, and to identify effective BCT clusters.

**Methods:** A systematic search of PubMed, Web of Science, and Embase was conducted from inception to December 18, 2023. Randomized and non-randomized controlled trials including adults with T2D were eligible if they evaluated behavior change or lifestyle interventions targeting PA, SB, and/or sleep and included at least one BCT. Data extraction, BCT coding (using the BCT Taxonomy), and risk of bias assessment (Cochrane RoB 2) were performed independently by multiple reviewers. Meta-analyses using random-effects models were conducted for relevant outcomes. Subgroup analyses examined the effects of three common BCT clusters: goals and planning, feedback and monitoring, and social support.

**Results:** Sixty-six studies (n = 18,725 participants) were included. Interventions significantly improved several PA outcomes, including steps/day (+1991 steps/day; p<0.001), total PA (SMD=0.36; p=0.02), moderate-to-vigorous PA (SMD=0.55; p<0.001), and light-intensity PA (SMD=0.62; p=0.01). Sedentary time decreased significantly (SMD=−0.32; p=0.008). Sleep quality improved (MD=−1.39; p=0.02), whereas sleep duration showed no significant change. Subgroup analyses demonstrated that BCT clusters involving goals and planning, feedback and monitoring, and social support were consistently associated with improvements in PA and SB, with comparable effect sizes to overall analyses. Effects on sleep outcomes were limited due to the small number of studies.

**Conclusion:** Behavior change interventions incorporating BCTs effectively increase PA, reduce SB, and improve sleep quality in adults with T2D. BCTs such as goal setting, self-monitoring, feedback, and social support appear particularly beneficial. However, sleep - especially duration - remains underexplored. Future interventions should adopt a 24-hour movement behavior perspective and more explicitly integrate and report BCTs to optimize long-term diabetes management.

## Introduction

Worldwide, diabetes is the 9^th^ leading cause of death, accounting for 6.7 million deaths in 2021 (1, 2). Over 90% of diagnoses are type 2 diabetes (T2D), a chronic disease characterized by hyperglycemia which eventually causes macro- (e.g., myocardial infarction, stroke) and microvascular complications (i.e., neuropathy, retinopathy, nephropathy) (1, 2). The global burden of T2D is rising, with prevalence forecast to increase to 783 million people by 2045 (1), posing significant challenges for the healthcare system. Next to a pharmacological approach, lifestyle has an important impact on the long-term management of diabetes (3). Improvements in for example physical activity (PA), sedentary behavior (SB), diet, stress and sleeping patterns are crucial for effective management of the disease (3). While lifestyle plays a crucial role in the management of T2D, adults with T2D tend to be less physically active and more sedentary compared with their peers (4).

A well-known paradigm shift emphasizes that three of these important lifestyle behaviors (i.e., PA, SB and sleep) are interrelated as part of a continuum of movement across one 24-hour day, the so-called 24-hour movement behaviors (24h-MBs). Every minute within a 24-hour time span can be categorized as either PA (subdivided into light-intensity PA (LPA) and moderate-to-vigorous PA (MVPA)), SB or sleep (5). As the daily sum of these behaviors is always 24-hour, they are by nature codependent and interchangeable. Changing the time spent on one behavior affects the duration of at least one of the other behaviors (5). Insights from observational studies using compositional data analysis, an innovative statistical approach for codependent data, demonstrate that theoretically replacing SB with either light-intensity or MVPA is associated with reductions in cardiometabolic risk (4, 6). For adults with T2D in particular, theoretically replacing SB with LPA was favorable for insulin, glucose and triglycerides, while MVPA was even more favorable (7). Moreover, the 24h-MB concept has recently been adopted by the International Diabetes Associations in their 2022 guidelines (8), emphasizing its recognized importance in counseling people with T2D.

As these guidelines and findings are primarily derived from observational studies and these guidelines need translations towards behavior change, it may be beneficial to develop interventions that target the composition of the entire day rather than focusing on single behaviors in isolation. However, changing individuals’ behaviors is inherently complex and presents numerous challenges (9). Therefore, intervention development should be firmly grounded in established theoretical frameworks such as the Theoretical Domains Framework (TDF) (10) or the COM-B model (11). Such frameworks offer a structured approach to understand behavioral determinants and provide guidance for selecting and applying appropriate behavior change techniques (BCTs). Selecting the correct BCTs is essential since they are the active components of interventions designed to modify these determinants, and a stronger theoretical foundation may thus enhance not only intervention effectiveness but also their uptake and long-term sustainability (12).

Several interventions already exist that apply BCTs to target specific health behaviors in adults with T2D. BCTs such as feedback and monitoring, shaping knowledge, goals and planning, and modifying antecedents are commonly used to improve lifestyle. A systematic review examining BCTs to improve PA among working adults with T2D found that psychologically informed educational approaches incorporating techniques such as goal setting, providing information about health consequences, demonstrating the behavior, and using prompts and cues were associated with improvements in both PA and glycemic control (13). Regarding SB, the use of wearable devices and app-based prompts appears to be promising in interrupting prolonged sitting among adults with T2D (14). Together, these findings indicate that theory-informed BCTs can effectively target individual behaviors relevant to T2D management. However, integrating such techniques within interventions that optimize 24-hour movement behaviors is not yet explored, and more specifically similarities or differences within clusters of BCTs across behaviors. Therefore, the aim of this systematic review and meta-analysis was to identify effective intervention components including BCTs that can positively change PA, SB and/or sleep in this population.

## Methods

This systematic review and meta-analysis was conducted following the Preferred Reporting Items for Systematic reviews and Meta-Analyses (PRISMA) guidelines (15). The protocol was registered on PROSPERO (ID: CRD42023494007).

### Search strategy

The literature was systematically searched to answer the following research question: ‘What is the effect of behavior change interventions using Behavior Change Techniques (Intervention) on PA, SB and/or sleep (outcome) in adults with T2D (population) compared to adults with T2D receiving usual care (comparison)?

Three electronic databases (PubMed, Web of Science and Embase) were searched from inception until 18/12/2023 using keywords of the following concepts: pathology (T2D), PA, SB, sleep, and study design (randomized controlled trials). The different search strategies for the three different electronic databases can be found in Additional file 1.

### Eligibility criteria

Covidence was used as an electronic library to manage the references from the final search strategy. Duplicates were first removed after which title and abstract were screened by paired independent reviewers (MDC & IW, LB & MK) using the Active Learning for Systematic Reviews (ASReview, version 1.3.4, Utrecht, the Netherlands). ASReview is an open-source artificial intelligence software that is designed to screen studies for systematic reviews. Before starting to screen, the active learning algorithm was trained by selecting relevant and irrelevant titles. The algorithm within the software then selects the most relevant studies for inclusion based on the initial training and the judgments made by the reviewers. Screening was stopped after 125 consecutive non-relevant studies appeared. Results after title and abstract screening were imported into Covidence where duplicates were removed. Full text screening was conducted by five independent reviewers (MDC, LB, IW, MK, MD). In addition, a manual search of the reference lists of included articles was conducted and additional articles were included if they complied with the in- and exclusion criteria.

The following inclusion criteria were used: (1) adults with T2D with a minimum age of 18 years old; (2) behavior change or lifestyle or multicomponent intervention(s); (3) randomized or non-randomized controlled trials; (4) the control group received usual care; (5) measurement of PA, SB and/or sleep; (6) inclusion of one or more BCT(s); (7) full text was available; and (8) articles written in English. The following exclusion criteria were used: (1) adults with other types of diabetes than T2D; (2) presence of comorbidities (e.g., T2D and neuropathy); (3) pilot or feasibility studies; (4) follow-up studies without control group; (5) the use of medication or nutritional/dietary supplements as an intervention; and (6) the control group receiving more than usual care (e.g., receiving a pedometer).

### Data extraction

Data were extracted by five independent reviewers (MDC, LB, IW, MK, MD). The following information was extracted (see additional file 2): (1) author and year of publication, (2) country, (3) study aims, (4) study design, (5) population, (6) number of groups, (7) sample size, (8) age, (9) sex, (10) description of the intervention, (11) targeted behaviors, (12) intervention duration, (13) intervention dose and frequency, (14) delivery method of the intervention, (15) which BCTs were included (either mentioned or assigned by the research team), (16) control group, (17) measurement method, (18) outcome, (19) means, standard deviations, 95% confidence intervals, and inter quartile ranges at pretest, posttest, follow-up.

Regarding BCTs, the BCT Taxonomy was applied to systematically identify and register the techniques used across the included studies (16). Three reviewers (LB, MDC, IW) independently coded the BCTs. Techniques were recorded when they were explicitly reported in the study. Additionally, BCTs were assigned based on detailed descriptions of the intervention content (Additional file 3). For example, the use of a pedometer to track step counts was coded as self-monitoring of behavior, even if not explicitly labeled as such by the original study authors.

### Risk of bias

The risk of bias was assessed by five independent reviewers (MDC, LB, IW, MK, MD) using the Cochrane Risk of Bias tool 2 (17). This checklist contains five different domains: (1) bias arising from the randomization process, (2) bias due to deviations from intended interventions, (3) bias due to missing outcome data, (4) bias in measurement of the outcome, and (5) bias in selection of the reported result. The Cochrane Risk of Bias tool 2 macro form was used to assess the risk of bias of all included articles. The macro contains an algorithm that produces a risk of bias judgement for each domain and was used to inform the assessor’s judgement. The algorithm provides a final risk of bias score (low risk, some concerns, high risk) which can be confirmed or changed by the assessor (Additional file 4).

### Data synthesis and analysis

Statistical analysis were conducted using Review Manager (RevMan) version 5.3. Based on the different outcomes identified in the studies, outcomes were categorized according to behavior. Outcomes reported for PA were steps/day, total PA (minutes/hours per day/week), Metabolic Equivalent of Task (MET)-minutes/hours, MVPA, LPA, moderate PA, vigorous PA, exercise (minutes or hours per day or week), walking (minutes or hours per day or week), counts/day, and calories expended. The outcome for SB was total sedentary time expressed in minutes/hours per day/week. Sleep was evaluated as sleep duration (hours/night) and sleep quality (score on the Pittsburgh Sleep Quality Index). Sample sizes used in the meta-analysis were sample sizes at the end of the intervention. In case of several follow-up measurements, data of the first measurement following the intervention were included. In addition, sample sizes based on completer analysis were included. Mean differences (MD; i.e., when the same outcome and unit of measure was used) and standardized mean differences (SMD; i.e., when the same outcome was used but different measures) with 95% confidence intervals (CI) were calculated to compare the effect of interventions between intervention group and control group for each outcome. The effects were visualized using forest plots. Based on the formula provided by Cochrane, means and standard deviations of intervention groups were combined when more than two intervention groups were compared against a control group (18). In addition, subgroup analyses were conducted based on the three most reported clusters of BCTs: goals and planning, feedback and monitoring, and social support. The effect size of the MDs and SMDs was evaluated using thresholds provided by Cohen (19): <0.20 = negligible, 0.20-0.49 = small, 0.50-0.79 = medium, and ≥0.80 = large. If a minimum of two studies evaluated the same outcome, pooled MDs or SMDs were calculated using a random-effects model using the inverse variance method. Heterogeneity of the included studies was assessed using I^2^ statistics. In case of high heterogeneity (I^2^ > 75%), a sensitivity analysis was performed by omitting studies with a high risk of bias. P-values <0.05 were considered significant.

## Results

In total, 9,437 were identified through the search strategy after which 66 studies eventually were included in this systematic review. The flow of the different processes (i.e, identification and screening) can be found in Figure 1.

**Figure 1.**
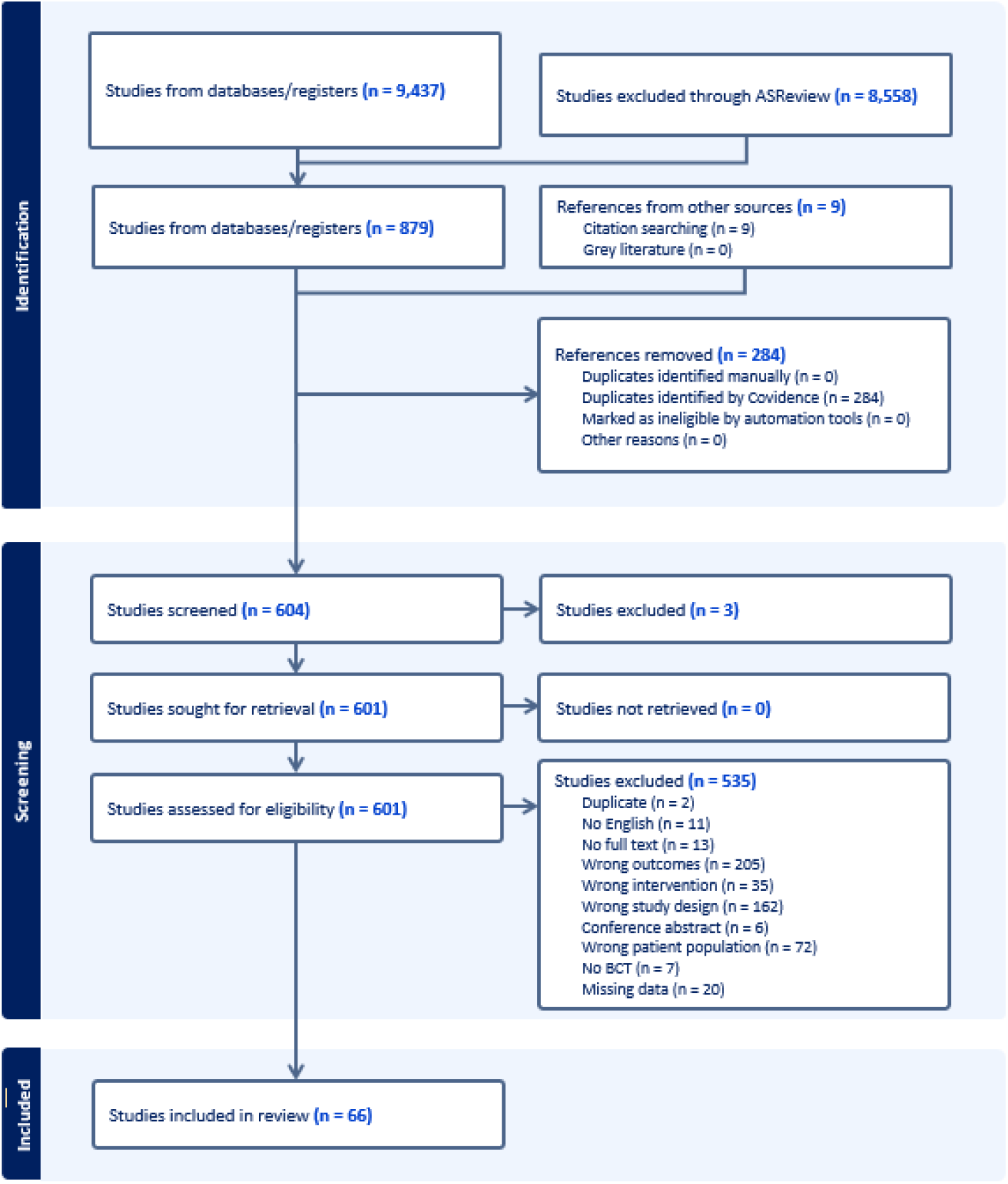
PRISMA Flowchart visualizing the flow of the identification and screening process.

The majority of the studies (n=62, 93.9%) were published between 2005 and 2025. Thirty-three percent (n=22) of the studies were conducted in North America, 28.8% (n=19) in Europe and 28.8% (n=19) in Asia. Only two studies (3.03%) were conducted in Africa and four (6.06%) in Oceania. In total, 18,725 participants were included across the different studies with 9,925 participants in the intervention group. Participants were 56.4 years old and 52.9% of them were female. The majority of the included studies (n=28, 42.4%) investigated total PA in isolation. Six studies (9.1%) investigated sleep in isolation and four studies (6.1%) focused on exercise. Almost all the other studies investigated two or more outcomes with a combination of total PA and MVPA as the most frequent studied combination (n=5, 7.6%) (Table 1).

**Table 1.** Study characteristics of included studies.

| Author et al.<br>year | Country | Study design | Number of<br>groups | Sample size<br>*At enrolment* |  | Age<br>(mean (SD))<br>*(median (IQR)) |  |  | Sex<br>(N and % female total sample) |  |  | Outcome<br>Movement<br>behavior(s)<br>(*sub*<br>*behaviors(s)*) | Instrument<br>(*Unit(s)*) |
| --- | --- | --- | --- | --- | --- | --- | --- | --- | --- | --- | --- | --- | --- |
|  |  |  |  | CG | IG | Total | CG | IG | Total | CG | IG |  |  |
| Akinci *et al.*<br>2018 | Turkey | single blind<br>RCT | 1. Supervised<br>exercise IG<br>2. Internet<br>based<br>exercise IG<br>3. CG | 22 | 22 | NR | 53.6<br>(6.7) | 1. 53.6<br>(6.0)<br>2. 50.2<br>(6.5) | NR | 15<br>(68.1) | 1. 18 (81.8)<br>2. 14 (66.6) | PA<br>(*steps/day*) | Pedometer<br>(*average steps*) |
| Alghafri *et al.*<br>2018 | Oman | Cluster RCT | 1. IG<br>2. CG | 110 | 117 | 44.2<br>(8.1) | 45.1<br>(9.2) | 43.5<br>(7.1) | 137<br>(59.1) | 71<br>(64.5) | 66 (54.1) | PA (*steps/day*,<br>*total PA -*<br>*METs*) | • Global Physical<br>ACGivity<br>Questionnaire (*MET*<br>*min/week*)<br>• ACGivpall (*steps/day*) |
| Alonso-Domí-<br>nguez *et al.*<br>2019 | Spain | RCT | 1. IG<br>2. CG | 102 | 102 | 60.6 | 60.4<br>(8.4)* | 60.8<br>(7.8)* | 92<br>(45.6) | 41<br>(40.2) | 52 (51.1) | PA (*steps/day*,<br>*total PA -*<br>*METs*) | • Pedometer<br>(*steps/day*)<br>• IPAQ (*total MET*<br>*min/week, MET min*<br>*MPA*) |
| Andrews et al.<br>2011 | UK | RCT | 1. Diet IG<br>2. Diet + PA IG<br>3. CG | 99 | 1. 24<br>8<br>246 | NR | 59.5<br>(11.1) | 1. 60.1<br>(10.2)<br>60.0 (9.7) | NR | 37<br>(37) | 1. 90 (36.0)<br>81 (34) | PA (*MVPA*) &<br>SB | • Digiwalker (*min/day*) |
| Araiza *et al.*<br>2006 | Mexico | RCT | 1. IG<br>2. CG | 15 | 15 | NR | 51 (10) | 49 (11) | NR | NR | NR | PA (*steps/day*) | Pedometer (*steps/day*) |
| Balducci *et al.*<br>2017 | Italy | RCT | 1. Behavioral IG<br>2. CG | 150 | 150 | NR | 62.3 (10.1) | 61.0 (9.7) | NR | 57 (38.0) | 59 (39.3) | PA (MVPA, LPA) & SB | Accelerometer<br>(*MET hours/week, min/day, hours/day*) |
| Balducci *et al.*<br>2019 | Italy | open-label, assessor-blinded RCT | 1. Behavioral IG<br>2. CG | 150 | 150 | NR | 62.3 (10.1) | 61.0 (9.7) | NR | 57 (38.0) | 59 (39.3) | PA (*total PA - METs, MVPA, LPA*) & SB | Accelerometer<br>(*MET hours/week, min/day, hours/day*) |
| Beverly *et al.*<br>2013 | USA | RCT | 1. IG<br>2. CG | 67 | 68 | 59.1 (8.7) | 58.4 (9.0) | 59.9 (8.5) | (51.5) | (55.2) | (47.8) | PA (*steps/day*) | Pedometer (*steps/day*) |
| Bjorgaas *et al.*<br>2005 | Norway | RCT | 1. IG<br>2. CG | 14 | 15 | 57.4 (7.8) | 56.9 (7.8) | 57.9 (8.0) | 0 (0) | 0 (0) | 0 (0) | PA (*steps/day*) | Pedometer (*steps/day*) |
| Cassidy *et al.*<br>2023 | UK | open-label, cluster-RCT | 1. IG<br>2. CG | 104 | 66 | NR | 57.0 (6.3) | 55.8 (6.6) | NR | 39 (37) | 30 (45) | PA (MVPA, LPA) & Sleep duration | Accelerometer<br>(*min/day*) |
| Christian *et al.*<br>2008 | Colorado (USA) | RCT | 1. IG<br>2. CG | 155 | 155 | NR | 53.4 | 53.0 | NR | 105 (33.9) | 100 (32.2) | PA (*total PA - METs*) | 7-day PA recall instrument (*Met min/week*) |
| De Greef *et al.*<br>2010 | Belgium | RCT | 1. IG<br>2. CG | 21 | 20 | NR | 61.3 (NR) | 61.3 (NR) | 13 (31.7) | 6 (28.6) | 7 (35) | PA (*steps/day, total PA – volume, MVPA, LPA*) & | • Yamax Digiwalker pedometer (*steps/day*) |
|  |  |  |  |  |  |  |  |  |  |  |  | SB |  |
| De Greef et al.<br>2011a | Belgium | RCT | 1. IG 1<br>2. IG 2<br>3. CG | 32 | 60 | 62<br>(9.0) | NR | NR | 29<br>(31) | NR | NR | PA (*steps/day*,<br>*total PA – volume, LPA*)<br>& SB | • Pedometer (*steps/day*)<br>• Accelerometer (*min/day*)<br>• IPAQ (*min/day*) |
| De Greef et al.<br>2011b | Belgium | RCT | 1. IG individual counseling<br>2. Intervnetio n group counseling<br>3. CG | 24 | 1. 22<br>2. 21 | 67.4<br>(9.3) | 66.0<br>(11.1) | 1. 66.6<br>(9.5)<br>70.0 (6.3) | 20<br>(29.9) | 7<br>(29.2) | 1. 5 (22.7)<br>8 (38.1) | PA (*steps/day*,<br>*total PA – volume, MVPA*) | • Yamax Digiwalker pedometer (*steps/day*)<br>• Diary (*min/day*)<br>• IPAQ long (*min/day*) |
| Dyson et al.<br>2010 | UK | RCT | 1. Video education IG<br>2. CG | 21 | 21 | 60.8<br>(9.6) | 62.9<br>(9.5) | 58.6 (9.2) | 24<br>(57.1) | 13<br>(61.9) | 11 (52.4) | PA (*steps/day*) | Pedometer (*steps/day*) |
| Eakin et al.<br>2013 | Australia | RCT | 1. Telephone counseling IG<br>2. CG | 151 | 151 | 58.0<br>(8.6) | 58.3<br>(9.0) | 57.7 (8.1) | 132<br>(43.7) | 65<br>(43) | 67 (44.4) | PA (*MVPA*) | ACGigraph accelerometer (*min/day*) |
| Fayehun et al.<br>2018 | Nigeria | RCT | 1. IG<br>2. CG | 23 | 23 | <40<br>years:<br>4.3%<br>(n=2)<br>40-59<br>years:<br>69.6<br>%<br>(n=32<br>) 60-64<br>years:<br>26.1<br>%<br>(n=12<br>) | <40<br>years:<br>4.3%<br>(n=1)<br>40-59<br>years:<br>69.6%<br>(n=16)<br>60-64<br>years:<br>26.1%<br>(n=6) | <40<br>years:<br>4.3%<br>(n=1)<br>40-59<br>years:<br>69.6%<br>(n=16)<br>60-64<br>years:<br>26.1%<br>(n=6) | 29<br>(63.0) | 15<br>(65.2) | 14 (60.9) | PA (*steps/day*) | Yamax pedometer<br>(*steps/day*) |
| Fritz et al.<br>2006 | Sweden | Non RCT | 1. IG<br>2. CG | 26 | 26 | NR | 59.3<br>(6.2) | 60.0 (7.3) | NR | 11<br>(42.3) | 15 (57.7) | PA (*exercise*) | PA Questionnaire<br>(*hours/week*) |
| Glasgow et al.<br>2010 | USA | RCT | 1. Computer<br>assisted self-management<br>(CASM) IG<br>2. CASM +<br>social<br>support IG<br>3. CG | 132 | 1. 16<br>9<br>162 | 58.4<br>(9.2) | 58.7<br>(9.1) | 1. 58.7<br>(9.3)<br>57.8 (9.3) | 231<br>(49.8) | 68<br>(51.5) | 1. 75 (44.6)<br>87 (53.7) | PA (*total PA-*<br>*calories,*<br>*exercise*) | CHAMPS (*kcal/day*) |
| Gray et al.<br>2021 | USA | RCT | 1. IG<br>2. CG | 142 | 145 | NR | 51.8<br>(9.4) | 53.5 (8.9) | NR | 71<br>(53.4) | 58 (44.6) | PA (*total PA -*<br>*volume*) | IPAQ (*min/week*) |
| Hashim et al.<br>2021 | Iraq | RCT | 1. IG<br>2. CG | 104 | 104 | NR | 46.5<br>(10.5) | 51.3<br>(11.6) | 130<br>(62.5) | 64<br>(61.5) | 66 (63.5) | PA (*total PA - METs*) | IPAQ (*min/week*) |
| Hirosaki et al.<br>2023 | Japan | RCT | 1. Laughter<br>yoga IG<br>2. CG | 21 | 21 | NR | 71.8<br>(6.4) | 70.6 (8.2) | NR | 14<br>(66.7) | 14 (66.7) | Sleep duration | Questionnaire<br>(*hours/night*) |
| Hordern et al.<br>2009 | Australia | RCT | 1. IG<br>2. CG | 112 | 111 | NR | 55 (8.5) | 56.1<br>(11.7) | NR | 48<br>(43) | 52 (47) | PA (*MPA, walking, VPA*) | Questionnaire<br>(*min/week*) |
| Irwig et al.<br>2012 | USA | RCT | 1. IG<br>2. CG | 51 | 52 | NR | 57.2<br>(9.7) | 54.8 (9.2) | NR | 29<br>(57) | 26 (50) | PA (*walking, exercise*) | Questionnaire<br>(*min/week*) |
| Jakicic et al.<br>2009 | USA | RCT | 1. IG<br>2. CG | 2575 | 2570 | 58.7<br>(6.8) | 58.8<br>(6.9) | 58.6 (6.8) | 2582<br>(50.2) | 1259<br>(48.9) | 1323 (51.5) | PA (*total PA-calories*) | Questionnaire<br>(*kcal/week*) |
| Jayasree &<br>Stalin<br>2019 | India | RCT | 1. IG<br>2. CG | 25 | 25 | NR | 60 (10) | 61 (10.2) | NR | 15<br>(60) | 16 (64) | SB | Self-reported<br>(*min/week*) |
| Kattelman et al.<br>2009 | USA | RCT | 1. IG<br>2. CG | 57 | 57 | NR | NR | NR | 87<br>(76) | NR | NR | PA (*LPA, MPA, VPA*) | Cross-cultural ACGivity<br>Participation PA survey<br>(*min/day*) |
| Keyserling et al.<br>2002 | USA | RCT | 1. Clinical and<br>community<br>based IG<br>2. Clinical IG<br>3. CG | 67 | 1. 67<br>2. 66 | NR | 59.2 | 1. 58.5<br>2. 59.8 | NR | 67<br>(100) | 1. 67 (100)<br>2. 66 (100) | PA (*total PA-calories*) | Caltrac accelerometer<br>(*Kcal/day*) |
| Khosravan et al.<br>2015 | Iran | RCT | 1. IG<br>2. CG | 34 | 34 | 57.8<br>(9.4) | NR | NR | 46<br>(68) | NR | NR | Sleep quality | PSQI (*score on 21*) |
| Kim et al.<br>2006 | Korea | RCT | 1. Web based IG<br>2. Paper based IG<br>3. CG | 23 | 1. 28<br>2. 22 | 55.1<br>(7.4) | NR | NR | 34<br>(46.6) | NR | NR | PA (*total PA - METs*) | Self-developed questionnaire (*MET hours/week*) |
| King et al.<br>2006 | USA | RCT | 1. IG<br>2. CG | 161 | 174 | 61.5<br>(11.3) | 61.0<br>(11.0) | 61.9<br>(11.7) | (50.2) | (50) | (51.3) | PA (*total PA-calories*) | CHAMPS (*kcal/kg/hour*) |
| Kirk et al.<br>2009 | Scotland | RCT | 1. In person PA consultation IG<br>2. Written PA consultation IG<br>3. CG | 35 | 1. 47<br>2. 52 | NR | 59.2<br>(10.4) | 1. 60.9<br>(9.6)<br>2. 63.2<br>(10.6) | NR | 17<br>(48.6) | 1. 22 (46.8)<br>2. 30 (57.7) | PA (*MPA*) | • ACGigraph GT1M accelerometer (*counts per week, steps/day*)<br>• Seven-day recall (*minutes/week*) |
| Lari et al.<br>2018 | Iran | Quasi-experimental study | 1. IG<br>2. CG | 40 | 40 | NR | 49.1 | 47.4 | NR | 17<br>(47.2) | 15 (37.5) | PA (*total PA - METs*) | Seven-day recall (*MET/week*) |
| Lee et al.<br>2015 | Australia | RCT | 1. IG<br>2. CG | 151 | 151 | 58.0<br>(8.6) | 58.3<br>(9.0) | 57.7 (8.1) | 132<br>(43.7) | 65<br>(43.0) | 67 (44.4) | PA (*total PA – volume, MVPA, walking*) | • ACGive Australia Survey (*min/week*)<br>• United States National Health Interview Survey (*min/week*)<br>ACGiGraph GT1M accelerometer<br>(min/week) |
| Lehmann et al.<br>1995 | Switzerland | RCT | 1. IG<br>2. CG | 21 | 13 | 55.5<br>(9.8) | NR | NR | NR | NR | NR | PA (total PA - volume) | Interview (min/week) |
| Li et al.<br>2023 | China | RCT | 1. IG<br>2. CG | 148 | 133 | 49.9<br>(11.8) | 52.4<br>(10.8) | 47.7<br>(12.3) | 113<br>(40.2) | 57<br>(42.9) | 56 (37.8) | Sleep quality | PSQI (score on 21) |
| Liebreich et al.<br>2009 | Canada | RCT | 1. IG<br>2. CG | 24 | 25 | NR | 54.5<br>(10.8) | 53.7 (9.8) | 29<br>(59) | NR | NR | PA (total PA - METs) | Modified version of Godin Leisure-Time Exercise Questionnaire (MET/min/week, min/week) |
| Liu et al.<br>2012 | China | RCT | 1. IG<br>2. CG | 89 | 119 | NR | 62.5<br>(10.0) | 61.9 (9.8) | NR | 55<br>(61.8) | 74 (62.2) | PA (exercise) | Questionnaire (min/week) |
| Lorig et al.<br>2009 | USA | RCT | 1. IG<br>2. CG | 159 | 186 | NR | 65.4<br>(11.4) | 67.7<br>(11.9) | NR | 105<br>(66.2) | 116 (62.4) | PA (exercise) | PA Scale (min/week) |
| Lorig et al.<br>2010 | USA | RCT | 1. IG<br>2. CG | 50 | 60 | 54.3<br>(NR) | NR | NR | 84<br>(76) | NR | NR | PA (exercise) | PA scale (min/week) |
| Lynch et al.<br>2019 | USA | Single blind<br>RCT | 1. IG<br>2. CG | 105 | 106 | 55.0,<br>(10.3) | 54.8<br>(9.0) | 55.1<br>(11.5) | 148<br>(70.1) | 73<br>(69.5) | 75 (70.8) | PA (steps/day, MPA) & SB | Accelerometer (steps/day, min/week) |
| Ma et al.<br>2008 | USA | RCT | 1. Low-glycemic index dietary education IG<br>2. CG | 21 | 19 | 53.5<br>(8.4) | 51.0<br>(8.25) | 56.31<br>(7.85) | 21<br>(52.5) | 10<br>(47.6) | 11 (57.9) | PA (*total PA - volume*) | Seven-day recall<br>(*min/week*) |
| Mahdizadeh et al.<br>2013 | Iran | RCT | 1. IG<br>2. CG | 42 | 42 | 48.4<br>(5.7) | NR | NR | 82<br>(100) | NR | NR | PA (*total PA - volume*) | IPAQ (*min/week*) |
| McEwen et al.<br>2017 | USA | RCT | 1. IG<br>2. CG | 74 | 83 | 53.5<br>(9) | 53.4<br>(8.4) | 53.6 (9.6) | 102<br>(65) | 53<br>(71.6) | 49 (59) | PA (*total PA - METs, MPA, walking*) | IPAQ (*MET min/week*) |
| Miyamoto et al.<br>2017 | Japan | RCT | 1. Non-locomotive PA IG<br>2. Locomotive PA consultation IG<br>3. CG | 10 | 1. 12<br>2. 10 | NR | 60.2<br>(3) | 1. 60,<br>(3.1)<br>2. 61.7<br>(1.9) | NR | 2 (20) | 1. 3 (25)<br>2. 2 (22.2) | PA (*total*) & SB | HJA-350IT accelerometer (*MET hors/600 min, min/day*) |
| Morowatishari fabad et al.<br>2021 | Iran | RCT | 1. IG<br>2. CG | 63 | 3. 63 | NR | 25-39 years:3<br>(4.8)<br>40-49 years:9<br>(14.5)<br>50-59 years:4<br>4 (71)<br>60-65 years:6 | 25-39 years: 3<br>(4.8)<br>40-49 years :14<br>(22.2)<br>50-59 years :41<br>(65.1) | NR | 48<br>(78.7) | 49 (77.8) | PA (*total PA - METs*) | IPAQ (*MET min/week*) |
|  |  |  |  |  |  |  | (9.7) | 60-65<br>years: 5<br>(7.9) |  |  |  |  |  |
| Negri et al.<br>2010 | Italy | RCT | 1. IG<br>2. CG | 21 | 39 | NR | 65.7<br>(5.2) | 65.7 (4.9) | NR | NR | NR | PA (*total PA - METs*) | PA record (*MET hours/week*) |
| Nepper et al.<br>2019 | USA | Quasi-<br>experimenta<br>l design | 1. IG<br>2. CG | 39 | 40 | NR | 55.7<br>(12.2) | 58.0<br>(10.6) | NR | 26<br>(67.1) | 26 (65.3) | PA (*total PA - METs*) | Block Physical Activity<br>Screening (MET<br>*min/week*) |
| Piette et al.<br>2011 | USA | RCT | 1. IG<br>2. CG | 167 | 172 | 56.0<br>(10.1) | 56.0<br>(10.9) | 55.1 (9.4) | 150<br>(51.5) | 73<br>(50) | 74 (51) | PA (*steps/day*) | Pedometer (*steps/day*) |
| Plotnikoff et<br>al.<br>2013 | Canada | RCT | 1. Print<br>material IG<br>2. Print<br>material and<br>telephone<br>counselling<br>IG<br>3. CG | 94 | 1. 97<br>2. 96 | NR | 61.0<br>(11.7) | 1. 61.4,<br>(12.6)<br>2. 62.3<br>(11.1) | NR | 44<br>(46.8) | 1. 39 (40.6)<br>2. 48 (51.0) | PA (*MVPA*) | Godin Leisure Time<br>Exercise Questionnaire<br>(*times/week*) |
| Poppe et al.<br>2019 | Belgium | 2 RCT's | 1. PA IG<br>2. SB IG<br>3. CG | 18 | 1. 24<br>2. 12 | 62.7<br>(8.4) | 64.9<br>(8.6) | 1. 62.91<br>(7.2)<br>59.0 (9.5) | 20<br>(37) | 9 (50) | 1. 7 (29)<br>4 (33) | PA (*total PA - volume, MVPA, LPA, MPA*) & SB | • ACGiggraph accelerometer (*min/day*)<br>• IPAQ (*min/day*)<br>LASA (*min/day*) |
| Samuel-Hodge<br>et al. | USA | RCT | 1. IG<br>2. CG | 18 | 36 | 54<br>(NR) | 53 (NR) | 55 (NR) | 40<br>(74) | 13<br>(72) | 27 (75) | PA (*VPA*) | • Questionnaire (*min/day*) |
| 2017 |  |  |  |  |  |  |  |  |  |  |  |  |  |
| Serin & Saritas<br>2021 | Turkey | RCT | 1. IG<br>2. CG | 76 | 76 | NR | 53.5<br>(8.8) | 51.3 (9.5) | NR | NR | NR | PA (*steps/day*) | Pedometer (*steps/day*) |
| Sinclair et al.<br>2023 | Saudi-Arabia | RCT | 1. IG<br>2. CG | 32 | 32 | 44.1<br>(9.2) | 45.9<br>(8.5) | 42.1 (9.7) | 33<br>(51.6) | 17<br>(53.1) | 16 (50) | Sleep quality | PSQI (*score on 21*) |
| Sung & Bae<br>2012 | Korea | Randomized<br>and<br>stratified<br>experimental<br>design | 1. IG<br>2. CG | 18 | 22 | NR | 70.1<br>(3.6) | 70.2 (4.7) | NR | 11<br>(61.1) | 15 (68.2) | PA (*total PA-calories*) | ACGigraph<br>accelerometer<br>(*aCGivity counts,*<br>*counts/min,*<br>*kcal/day/kg*) |
| Thuita et al.<br>2020 | Kenya | RCT | 1. Nutrition<br>education IG<br>2. Nutrition<br>education<br>and peer to<br>peer support<br>intervention<br>3. CG | 51 | 1. 51<br>51 | NR | 56<br>(12.0) | 1. 55<br>(12.3)<br>57 (10.9) | NR | 30<br>(58.8) | 1. 27 (52.9)<br>34 (66.7) | PA (*total PA -*<br>*METs*) | PA questionnaire (*MET*<br>*min/week*) |
| Toobert et al.<br>2007 | USA | RCT | 1. IG<br>2. CG | 116 | 2. 16<br>3 | NR | 60.7<br>(7.8) | 61.1 (8.0) | (100) | (100) | (100) | PA (*total PA-calories*) | CHAMPS<br>(*kcal/kg/hour*) |
| Tudor-Locke<br>et al.<br>2004 | UK | RCT | 1. IG<br>2. CG | 30 | 30 | 52.7<br>(5.2) | 52.5<br>(4.8) | 52.8 (5.7) | 11<br>(23.4) | 9<br>(39.1) | 12 (50.0) | PA (*steps/day*) | Pedometer (*steps/day*) |
| Unick et al.<br>2016 | USA | RCT | 1. Intensive lifestyle IG<br>2. CG | 1201 | 1199 | 59.3<br>(6.9) | 59.5<br>(6.8) | 59.1 (6.9) | 1340<br>(55.8) | 684<br>(57.0) | 656 (54.7) | PA (*total PA - METs*) | Accelerometer (*MET min/week*) |
| Van Dyck et al.<br>2013 | Belgium | RCT | 1. IG<br>3. CG | 32 | 60 | 62<br>(9.0) | NR | NR | 29<br>(31.0) | NR | NR | PA (*steps/day*) | Pedometer (*steps/day*) |
| Varney et al.<br>2014 | Australia | RCT | 1. IG<br>2. CG | 47 | 47 | NR | 64 [61-66]<br>(mean [95% CI]) | 59 [56-62] | NR | 17<br>(36) | 13 (28) | PA (*total PA - volume*) | New-Zealand Physical Activity Questionnaire (*kJ/kg/day*) |
| Vluggen et al.<br>2021 | Netherlands | RCT | 1. IG<br>2. CG | 244 | 234 | NR | 59.4<br>(7.1) | 60.9 (6.3) | NR | 79<br>(32.4) | 76 (32.5) | PA (*total PA - volume*) | Short Questionnaire to Assess Health-Enhancing Physical Activity (*min/week*) |
| Welschen et al. 2013 | Netherlands | RCT | 1. IG<br>2. CG | 78 | 76 | NR | 61.2<br>(8.8) | 60.5 (9.4) | NR | 28<br>(35.8) | 31 (40.5) | PA (*LPA, MPA, VPA*) | Short Questionnaire to Assess Health-Enhancing Physical Activity (*min/day*) |
| Yoo et al.<br>2008 | Korea | RCT | 1. Continuous glucose monitoring IG<br>2. CG | 33 | 32 | NR | 57.5<br>(9.0) | 54.6 (6.8) | NR | 14<br>(50) | 19 (65.5) | PA (*exercise*) | Diary (*min/week*) |
| Zhang et al.<br>2021 | China | cluster RCT | 1. IG<br>2. CG | 568 | 574 | NR | 61.8<br>(8.6) | 61.6 (9.2) | NR | 383<br>(67.4) | 370 (64.5) | Sleep quality | PSQI (*Score on 21*) |
| Zuo et al.<br>2020 | China | single-blind<br>RCT | 1. IG<br>2. CG | 95 | 96 | NR | 61.7<br>(10.4) | 63.9<br>(10.2) | NR | 64<br>(68.8) | 62 (66.0) | Sleep quality | PSQI (*Score on 21*) |

### The effects of behavior change interventions on physical activity, sedentary behavior and sleep Physical activity

#### Steps/day

In total, 17 studies including 1,921 participants investigated the effect of behavior change interventions on steps/day in adults with T2D (**Figure 2**) (20–36). The pooled SMD in steps/day between the intervention and control groups was +1991 steps/day (CI: 1220-2763; p<0.001). This shows that behavior change interventions resulted in a statistically significant increase in daily steps. Sensitivity analyses excluding studies with high risk of bias showed that the results were consistent across subgroups, suggesting that heterogeneity is not related to quality of the included studies.

**Figure 2.**
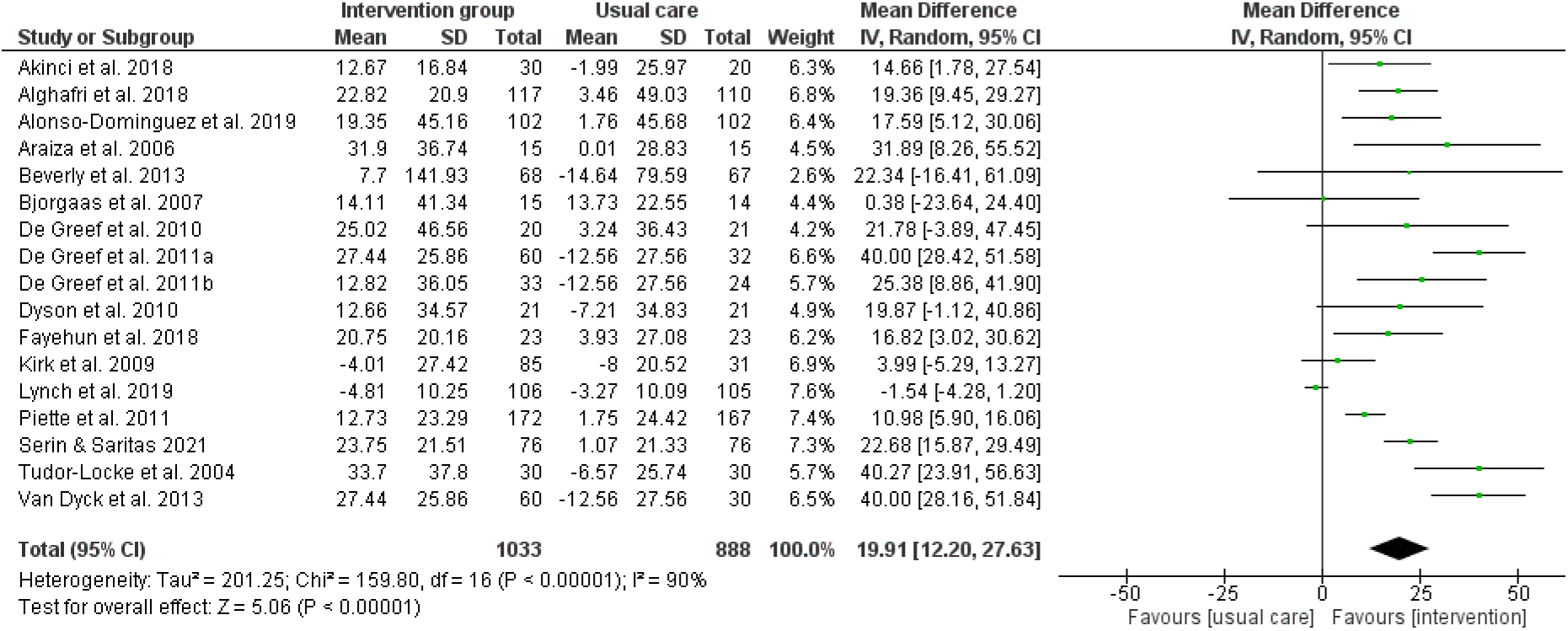
Forest plot of the effects on steps/day. Means and standard deviations should be multiplied by a factor 100 to reflect the total amount of steps/day.

#### Total physical activity

Total PA had three different outcome measures: volume, METs and calories. Eleven studies, including 1,285 participants, evaluated the effect of behavior change interventions on the volume (expressed in minutes/day or per week) of PA (**Figure 3.1**) (26–28, 37–44). The pooled SMD between the intervention and control groups was +0.36 (CI 0.05-0.66, p = 0.02), indicating significant increase in PA volume in the intervention group. The results remained the same after sensitivity analysis. Fourteen studies (n = 3,518) investigated the effects of a behavior change intervention on METs showing a SMD of +1.17 (CI −0.11– 2.45, p=0.07), which was not significant (**Figure 3.2**) (21, 22, 45–56). After sensitivity analysis, a significant SMD was found (+0.61; CI 0.30-0.91, p<0.001). Six studies (n=3,325) examined the differences in calorie expenditure after the implementation of a behavior change intervention (**Figure 3.3**) (57–62). The SMD between the intervention and control group was +0.55 (0.21-0.88, p=0.001). Significance remained after sensitivity analysis.

**Figure 3.**
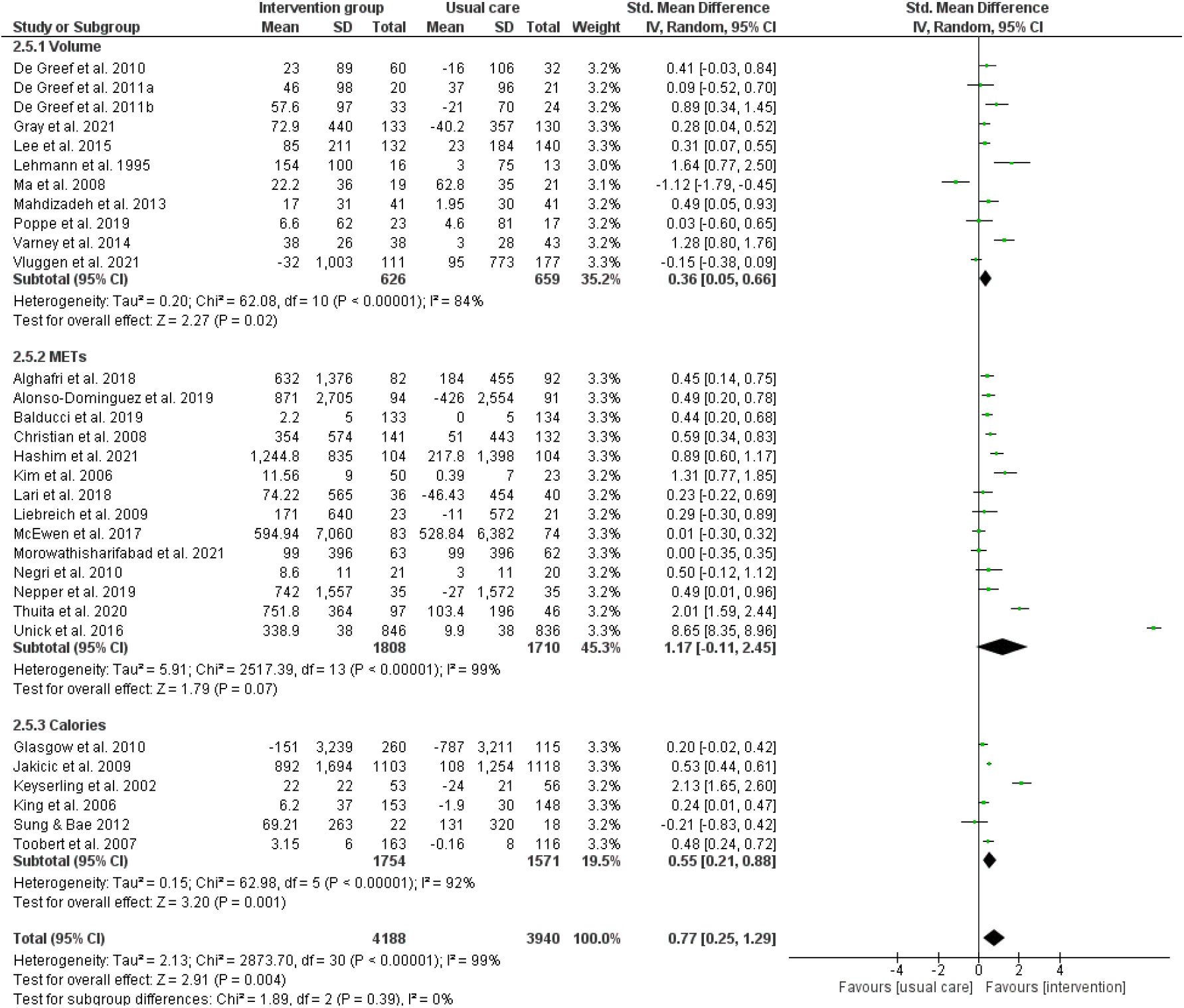
Forest plot of the effects on total physical activity.

#### Moderate-to-vigorous physical activity

Eleven studies including 2,391 participants evaluated the effect of behavior change interventions on MVPA (**Figure 4**) (26–28, 42, 45, 63–67). The pooled SMD between the intervention and control groups was +0.55 (CI: 0.26-0.84; p<0.001) reflecting a large effect size. Behavior change interventions resulted in a statistically significant increase in MVPA in adults with T2D. No sensitivity analysis was performed as no studies with high risk of bias were included.

**Figure 4.**
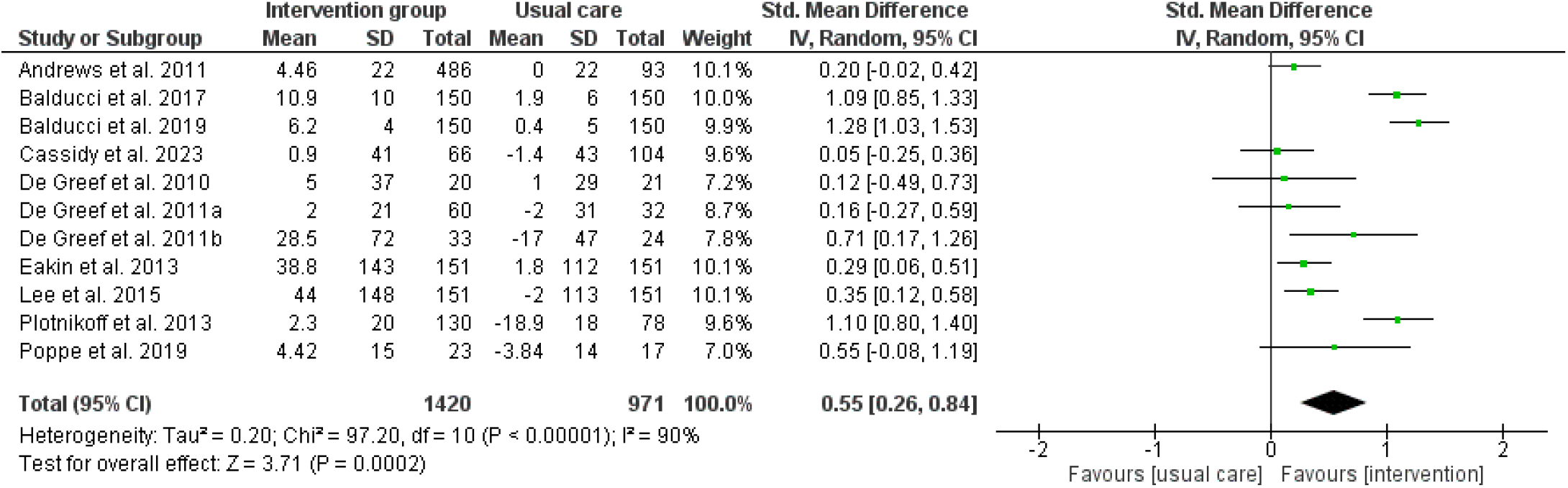
Forest plot of the effects on moderate-to-vigorous physical activity.

#### Light-intensity physical activity

Eight studies including 1,168 participants evaluated the effect of behavior change interventions on LPA (**Figure 5**) (26, 27, 42, 45, 64, 65, 68, 69). The pooled SMD between the intervention and control groups was +0.62 (CI: 0.13-1.11; p=0.01) reflecting a medium effect size. Behavior change interventions resulted in a medium statistically significant increase in LPA in adults with T2D. Sensitivity analyses showed similar results.

**Figure 5.**
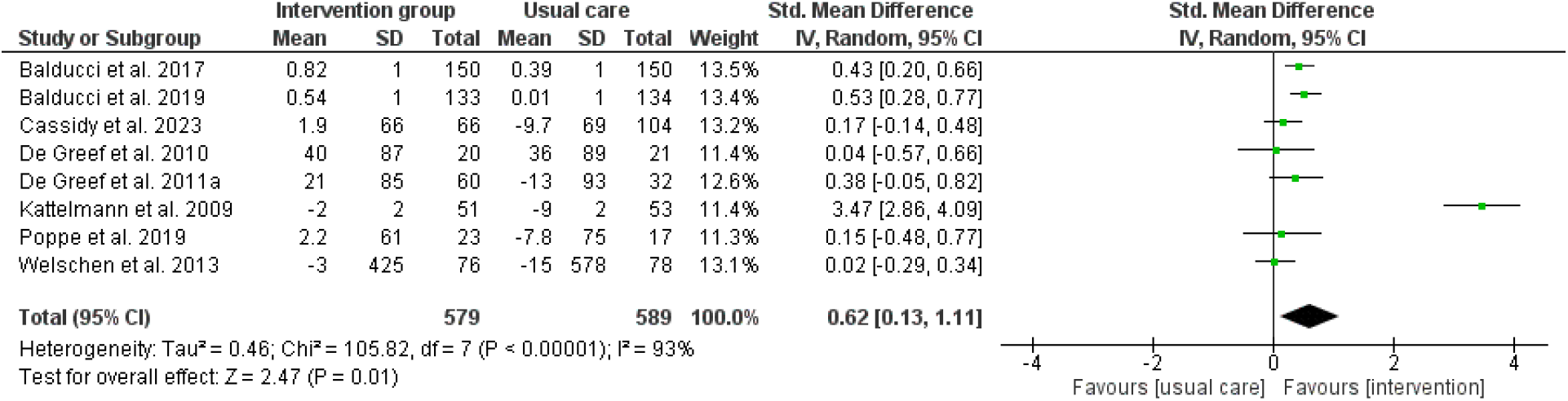
Forest plot of the effects on light-intensity physical activity.

#### Moderate physical activity

In total, seven studies (n = 957) evaluated the effect of behavior change interventions on moderate PA (**Figure 6**) (32, 33, 42, 51, 68–70). The pooled SMD between the intervention and control groups was +0.43 (CI: 0.03-0.84; p=0.04) reflecting a significant small increase in moderate PA in the intervention group. The results remained the same after sensitivity analysis.

**Figure 6.**
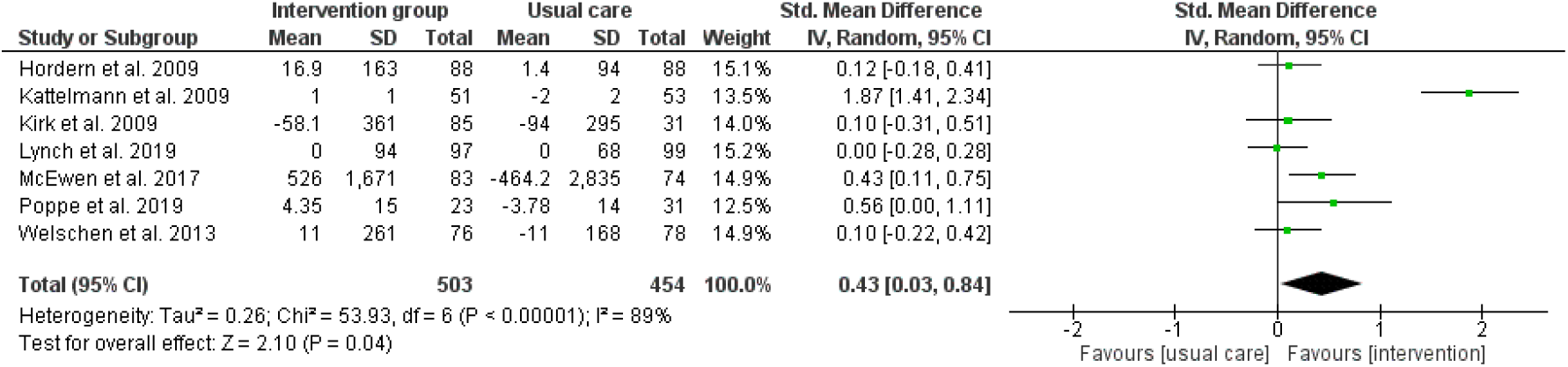
Forest plot of the effects on moderate physical activity.

#### Vigorous physical activity

Five studies including 645 participants evaluated the effect of behavior change interventions on vigorous PA (**Figure 7**) (51, 68–71). The pooled SMD between the intervention and control groups was 0.75 (0.07-1.42, p=0.03). Behavior change interventions resulted in a statistically significant increase in vigorous PA in adults with T2D. Sensitivity analysis showed no significant effect on vigorous PA.

**Figure 7.**
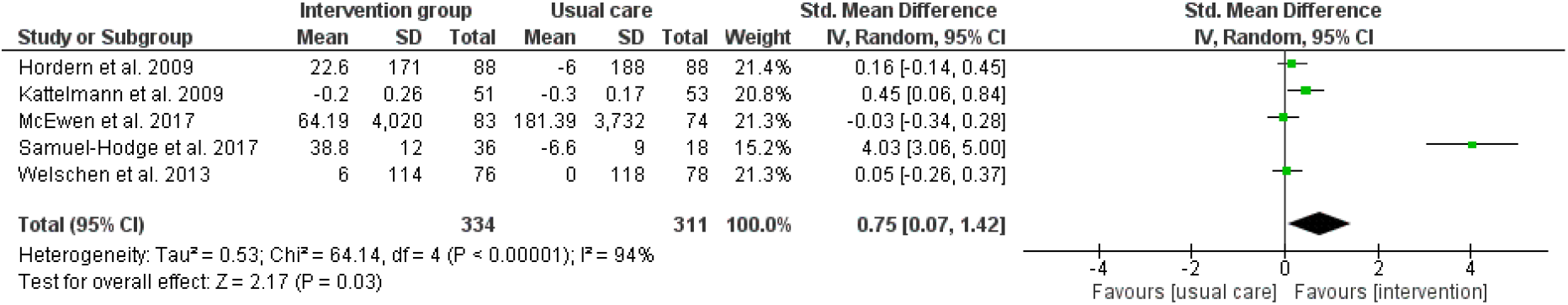
Forest plot of the effects on vigorous physical activity.

#### Exercise

Six studies (n = 740) evaluated the effect of behavior change interventions on exercise (**Figure 8**) (72–77). The pooled SMD between the intervention and control groups was +0.26 (CI: 0.06-0.46; p=0.01) reflecting a small but statistically significant increase in time spent on exercise in the intervention group of adults with T2D. Sensitivity analysis showed no significant effect on exercise.

**Figure 8.**
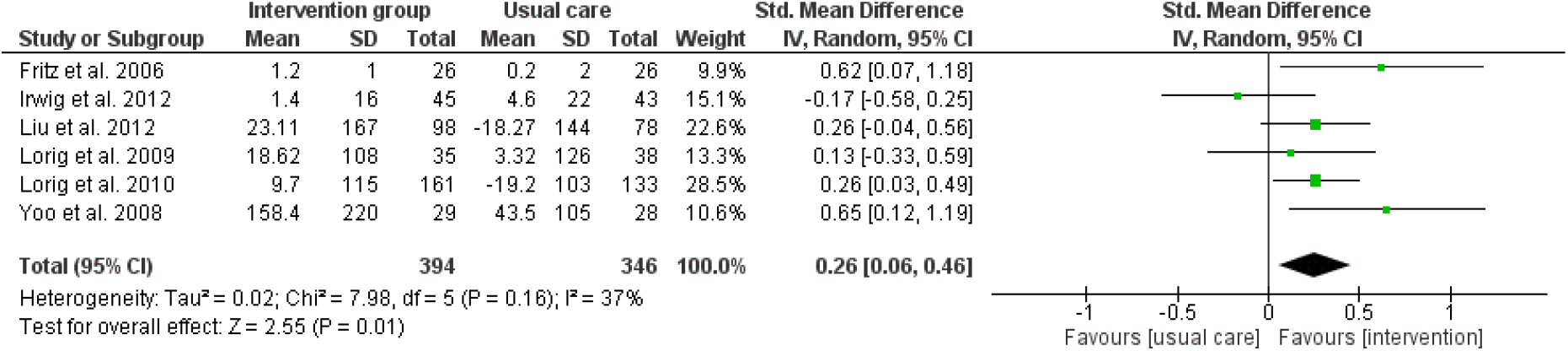
Forest plot of the effects on exercise.

#### Walking

Four studies including 805 participants evaluated the effect of behavior change interventions on walking (in minutes or hours/day or minutes or hours/week) (**Figure 9**) (38, 51, 70, 73). The pooled SMD between the intervention and control groups was 0.00 (CI: −0.18-0.19; p=0.99). Behavior change interventions did not lead to an increase in time spent walking in adults with T2D. Sensitivity analyses was not required.

**Figure 9.**
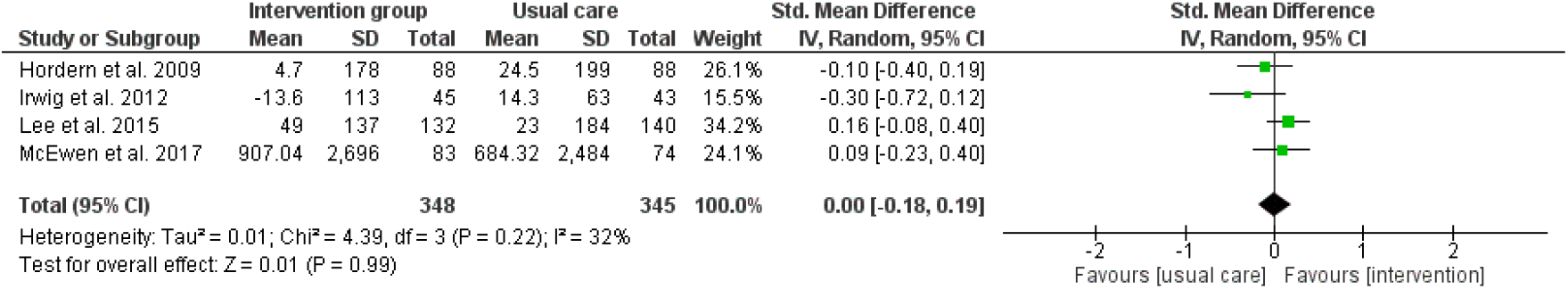
Forest plot of the effects on walking.

### Sedentary behavior

Nine studies (n=1,644) evaluated the effect of behavior change interventions on sedentary time (**Figure 10**) (26, 28, 33, 42, 45, 63, 64, 78, 79). The pooled SMD between the intervention and control groups was −0.32 (CI: −0.56, −0.008; p=0.008). Behavior change interventions were effective in decreasing sedentary time in adults with T2D. Sensitivity analyses was not required (I^2^=74%).

**Figure 10.**
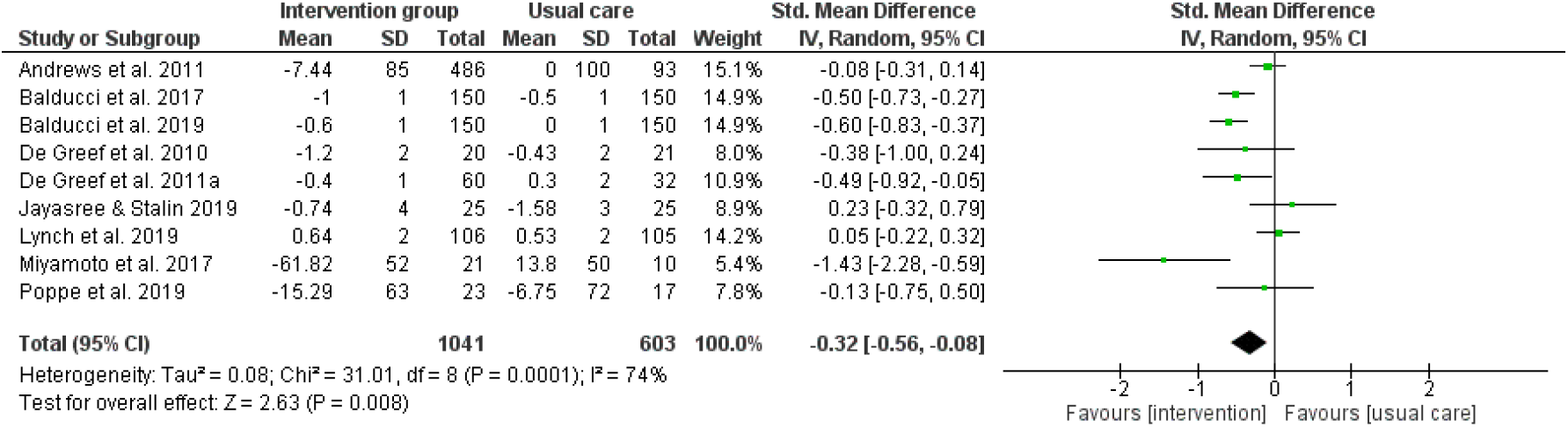
Forest plot of the effects on sedentary time.

### Sleep

#### Sleep duration

Only two studies including 212 participants evaluated the effect of behavior change interventions on sleep duration (**Figure 11**) (65, 80). The pooled mean standardized difference between the intervention and control groups was +0.25 (CI: −0.03-0.52; p=0.08). Behavior change interventions did not result in a significant increase in sleep duration in adults with T2D. The I^2^ value was 0% indicating low heterogeneity.

**Figure 11.**
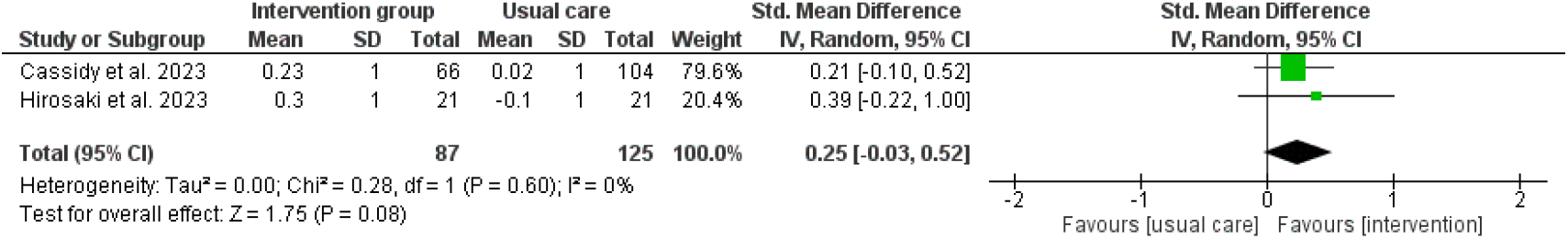
Forest plot of the effects on sleep duration.

#### Sleep quality

Five studies including 1,627 participants evaluated the effect of behavior change interventions on sleep quality measured with the Pittsburgh Sleep Quality Index (**Figure 12**) (81–85). The pooled SMD between the intervention and control groups was −1.39 (CI: −2.55—0.24; p=0.02). Behavior change interventions were effective in increasing sleep quality in adults with T2D. No studies with high risk of bias were included in the analysis.

**Figure 12.**
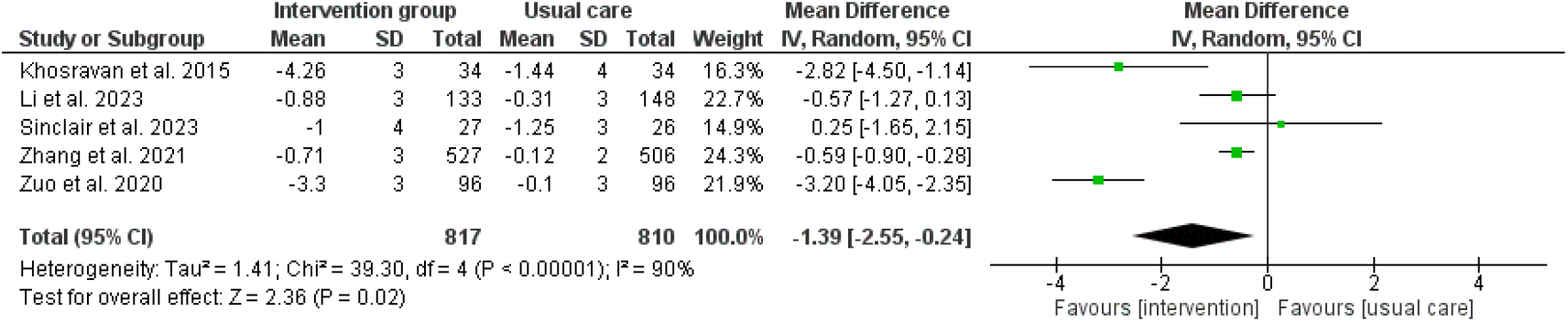
Forest plot of the effects on sleep quality.

### The effects of BCTs within the cluster of goals and planning

The results from the sub analysis on studies that reported one or more BCTs can be found below. Results are presented according to one of the three mostly used clusters of BCTs, namely goals and planning, feedback and monitoring, and social support. Analyses were conducted for the outcomes that were mostly studied: steps per day, total PA, MVPA, sedentary time, and sleep quality.

Most used BCTs within the cluster of goals and planning are: goal setting (behavior), problem solving, goal setting (outcome), action planning, and review behavior goals [see supplementary file x]. Overall, using BCTs from the cluster ‘goals and planning’ resulted in positive effects on steps/day (SMD +2,263 steps/day; CI 1156,3370; p<0.001), total PA (SMD +0.72; CI 0.13,1.30; p=0.02), MVPA (SMD +0.55; CI 0.26,0.84; p<0.001), and sedentary time (SMD −0.31; CI −0.53,-0.08; p=0.007). These results are comparable with the overall analyses and show that interventions using goals and planning in adults with T2D can lead to beneficial changes in PA and sedentary time. No studies on sleep quality included BCTs focusing on goals and planning.

**Figure 13.**
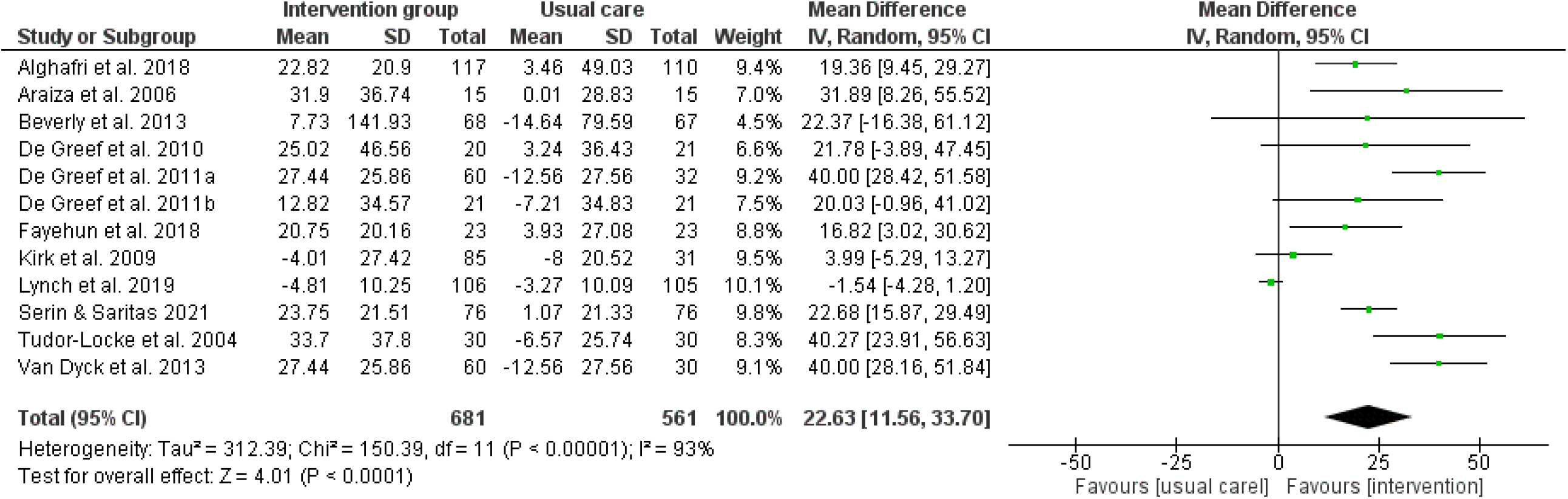
Forest plot of the effects of BCTs in the cluster ‘goals and planning’ on steps/day. Means and standard deviations should be multiplied by a factor 100 to reflect the total amount of steps/day.

**Figure 14.**
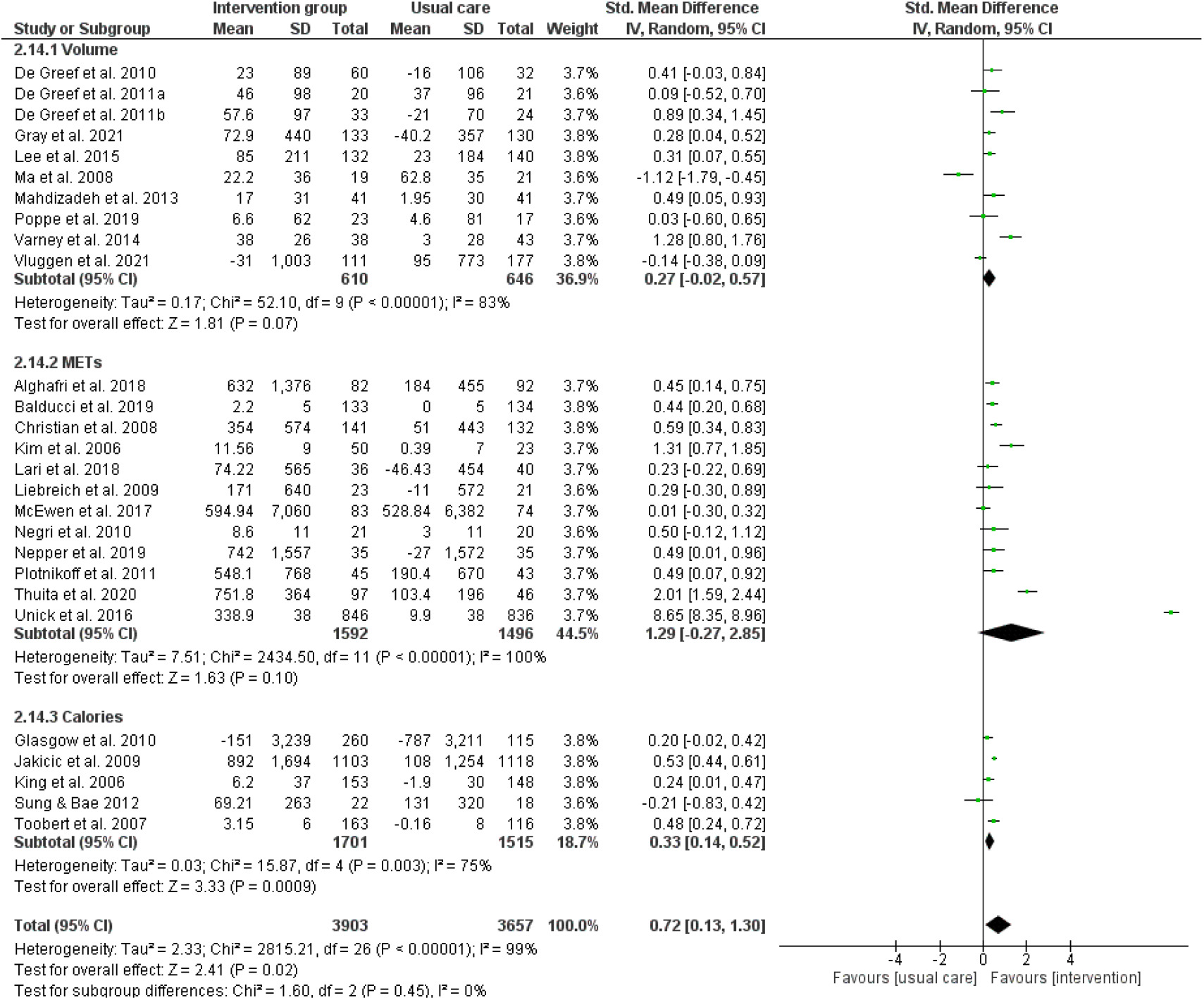
Forest plot of the effects of BCTs in the cluster ‘goals and planning’ on total PA.

**Figure 15.**
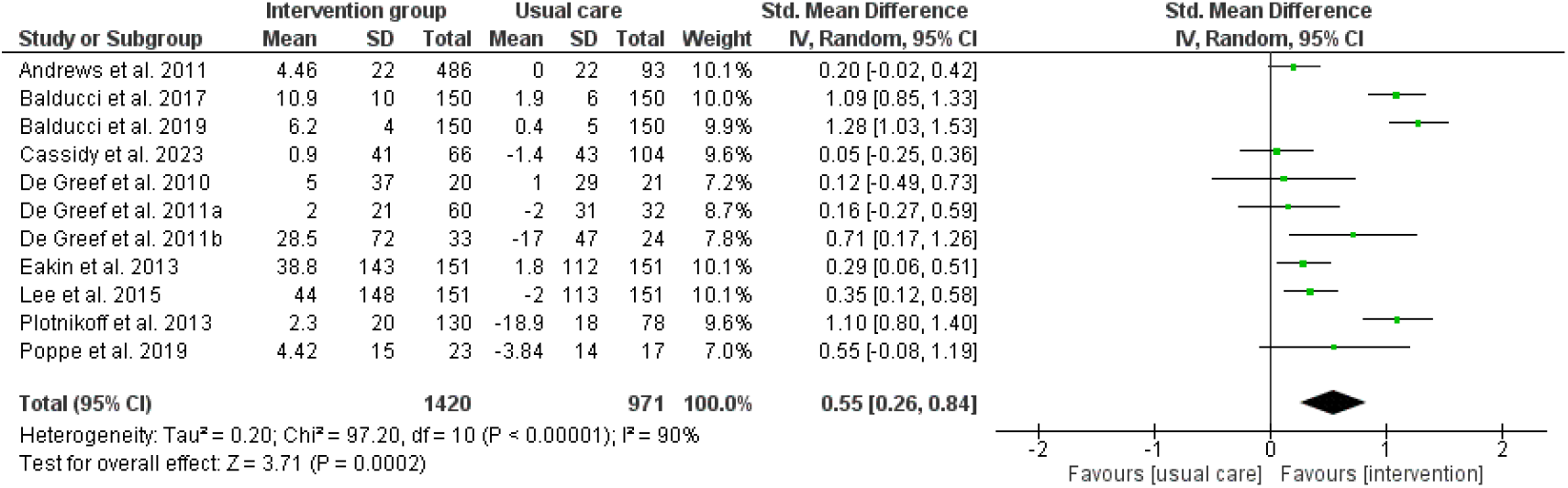
Forest plot of the effects of BCTs in the cluster ‘goals and planning’ on MVPA.

**Figure 16.**
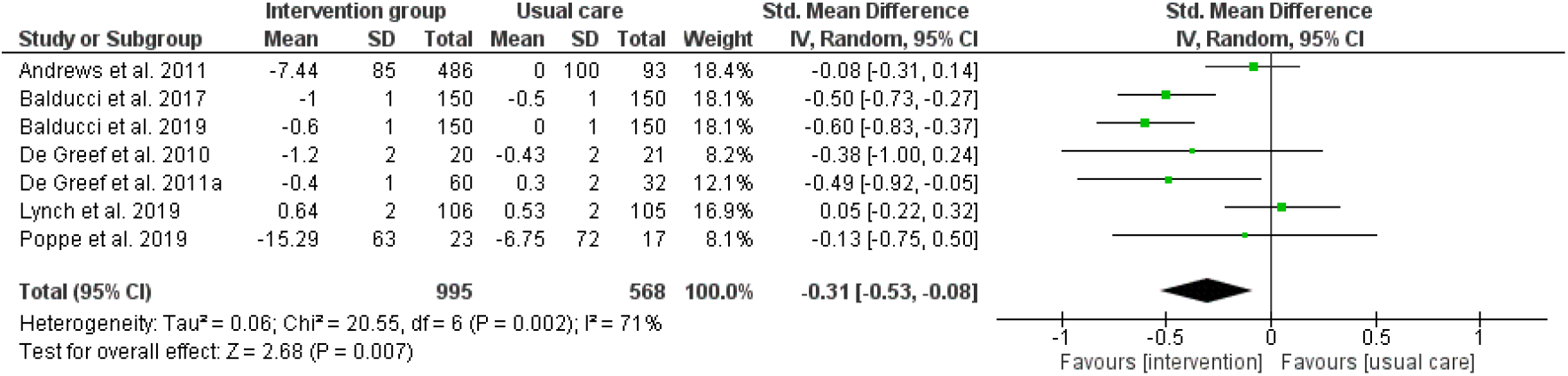
Forest plot of the effects of BCTs in the cluster ‘goals and planning’ on sedentary time.

### The effects of BCTs within the cluster of feedback and monitoring

Most used BCTs within the cluster of feedback and monitoring are: feedback on behavior, self-monitoring of behavior, self-monitoring of outcomes of behavior, feedback on outcomes of behavior [see supplementary file x]. Using BCTs from the cluster ‘feedback and monitoring’ resulted in positive effects on steps/day (n=13; SMD +1917 steps/day; CI 1106,2728; p<0.001), total PA (n=20; SMD +0.77; CI 0.02,1.51; p=0.04), MVPA (n=9; SMD +0.56; CI 0.26,0.86; p<0.001), and sedentary time (n=7; SMD − 0.36; CI −0.63,-0.09; p=0.009). No significant effects of feedback and monitoring were found for sleep quality (n=2). These results are comparable with the overall analyses and show that interventions using feedback and monitoring in adults with T2D can lead to beneficial changes in PA and sedentary time. However, these results were not significant for volume or METs when looking at total PA

**Figure 17.**
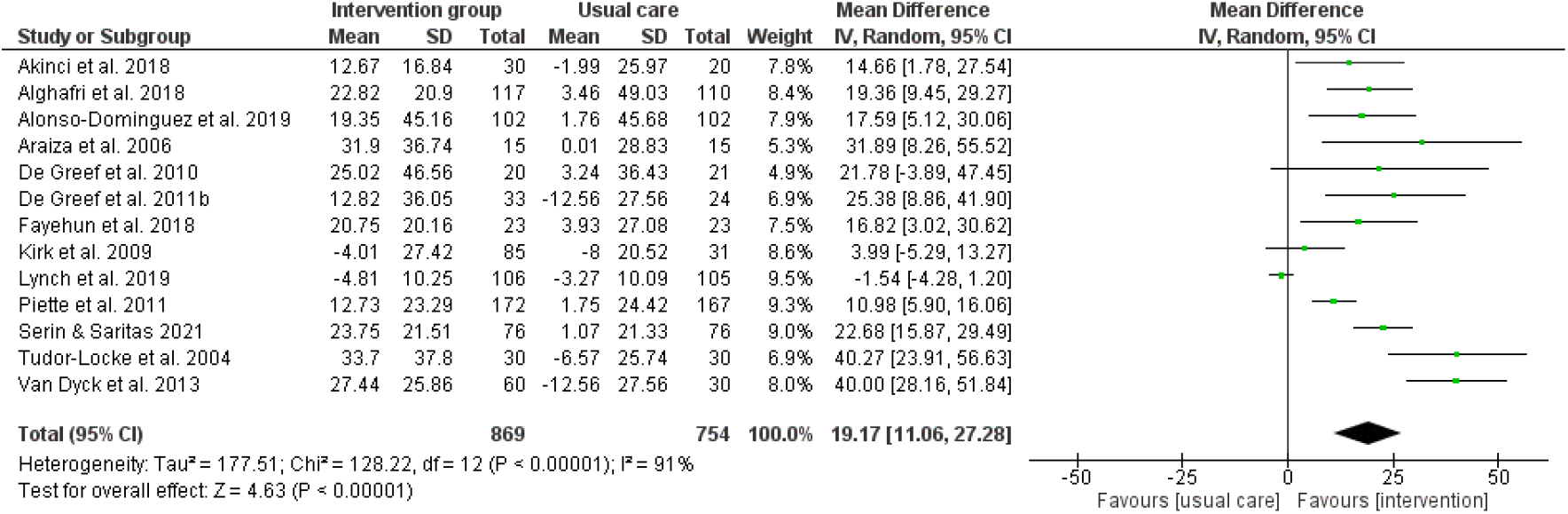
Forest plot of the effects of BCTs in the cluster ‘feedback and monitoring’ on steps/day. Means and standard deviations should be multiplied by a factor 100 to reflect the total amount of steps/day.

**Figure 18.**
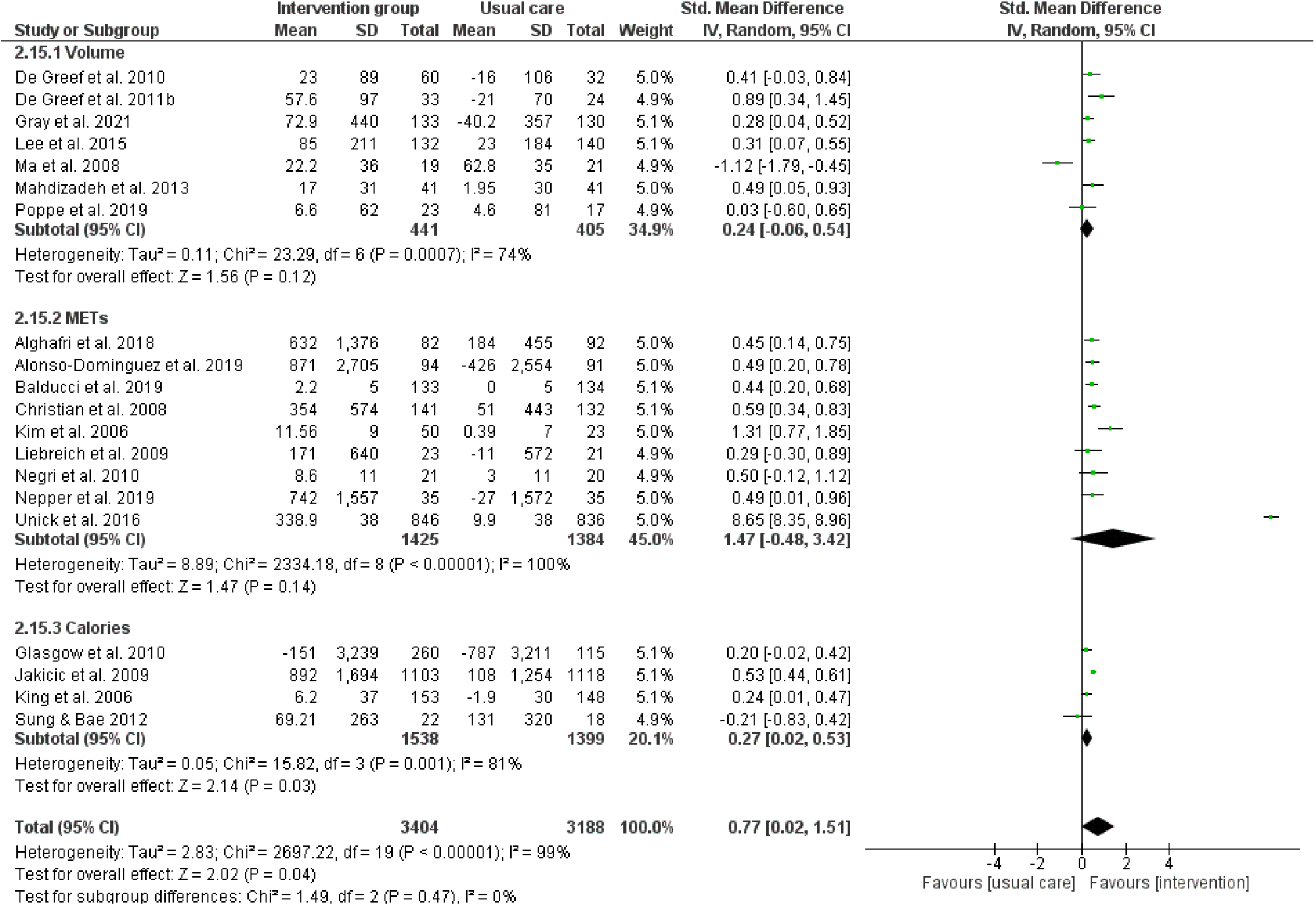
Forest plot of the effects of BCTs in the cluster ‘feedback and monitoring’ on total PA.

**Figure 19.**
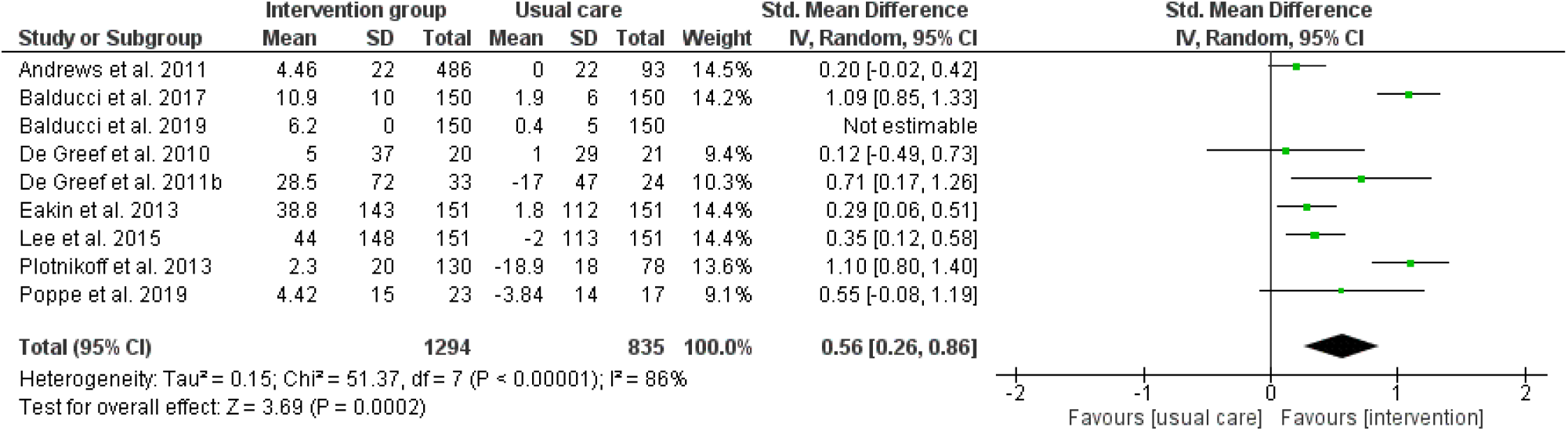
Forest plot of the effects of BCTs in the cluster ‘feedback and monitoring’ on MVPA.

**Figure 20.**
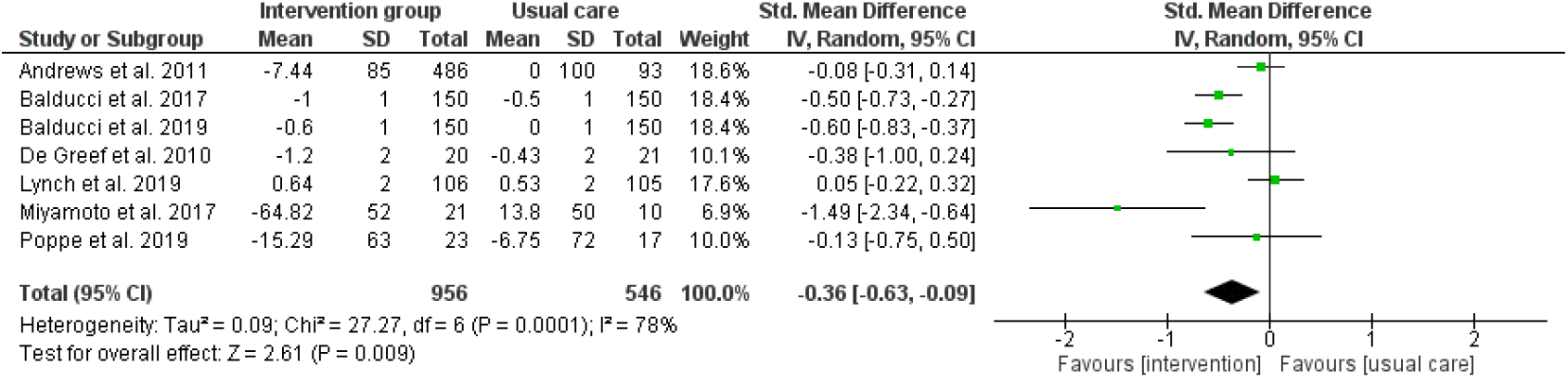
Forest plot of the effects of BCTs in the cluster ‘feedback and monitoring’ on sedentary time.

**Figure 21.**
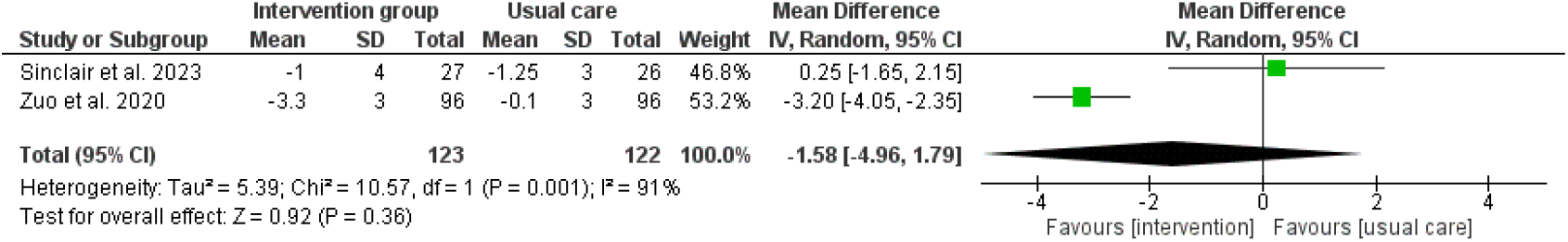
Forest plot of the effects of BCTs in the cluster ‘feedback and monitoring’ on sleep quality.

### The effects of BCTs within the cluster of social support

Using BCTs from the cluster ‘social support’ resulted in positive effects on steps/day (n=9; SMD +2059 steps/day; CI 758,3360; p=0.002), MVPA (n=4; SMD +0.53; CI 0.06,1.00; p=0.03), and sedentary time (n=6; SMD −0.36; CI −0.60,-0.11; p=0.005). These results are comparable with the overall analyses and show that interventions using social support in adults with T2D can lead to beneficial changes in PA and sedentary time. However, these results were not significant for total PA. No studies on sleep quality included BCTs focusing on social support. Most used BCT within the cluster of social support was social support (unspecified) [see supplementary file x].

**Figure 22.**
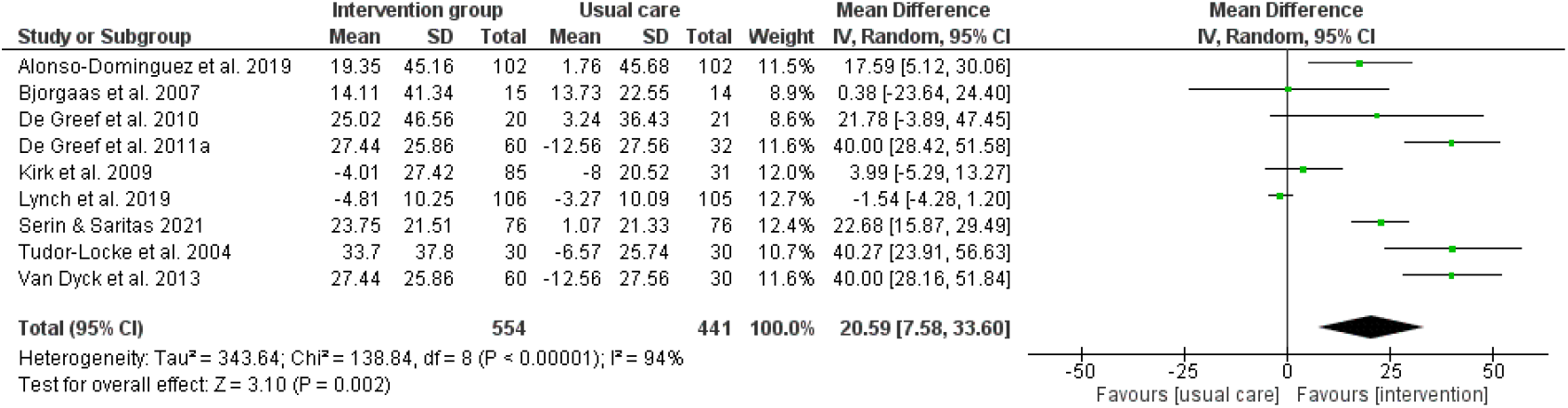
Forest plot of the effects of BCTs in the cluster ‘social support’ on steps/day. Means and standard deviations should be multiplied by a factor 100 to reflect the total amount of steps/day.

**Figure 23.**
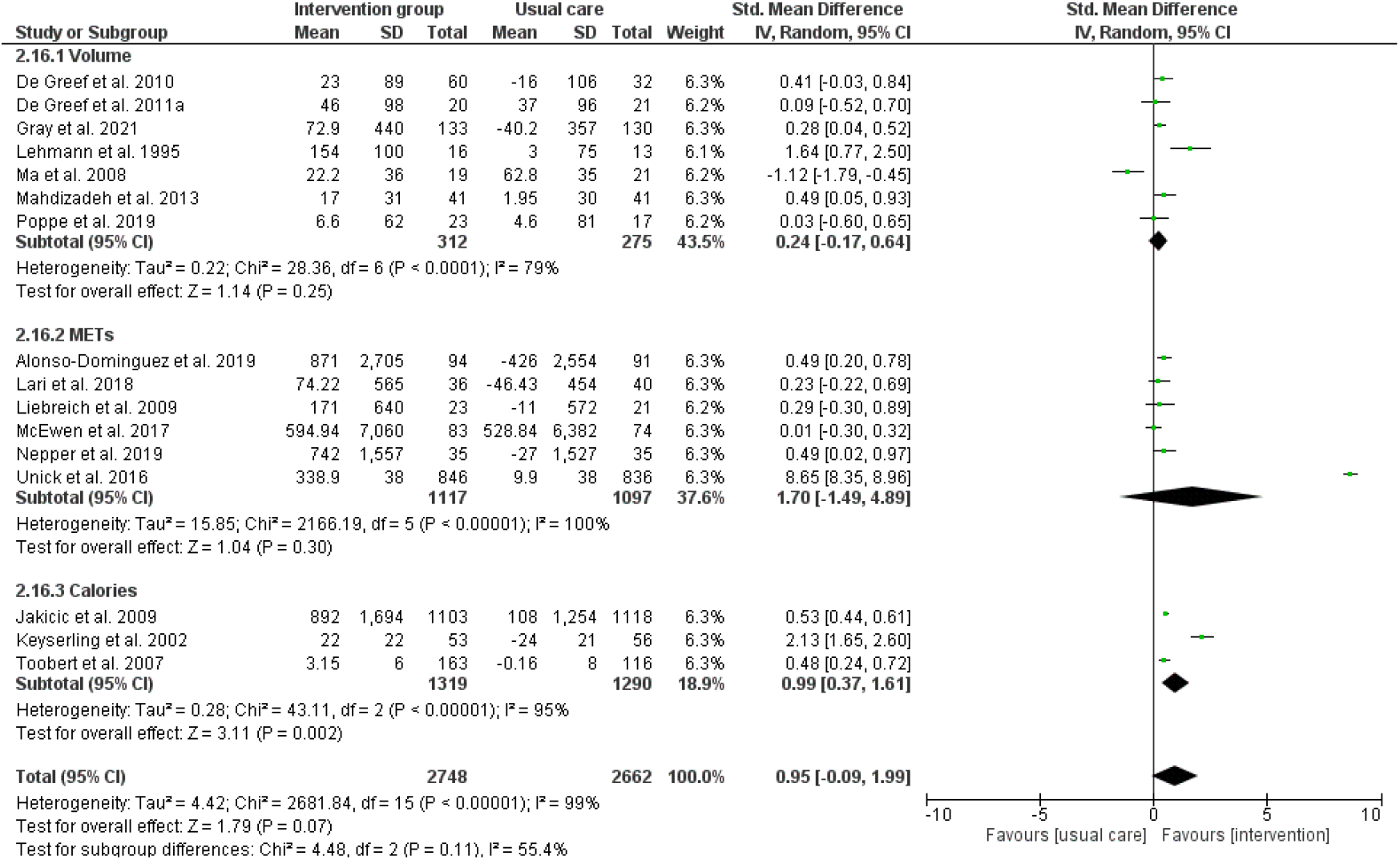
Forest plot of the effects of BCTs in the cluster ‘social support’ on total PA.

**Figure 24.**
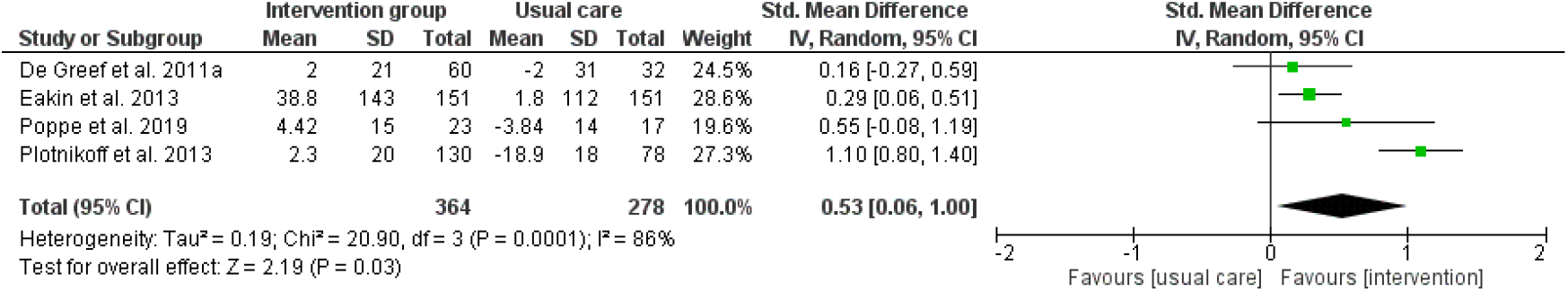
Forest plot of the effects of BCTs in the cluster ‘social support’ on MVPA.

**Figure 25.**
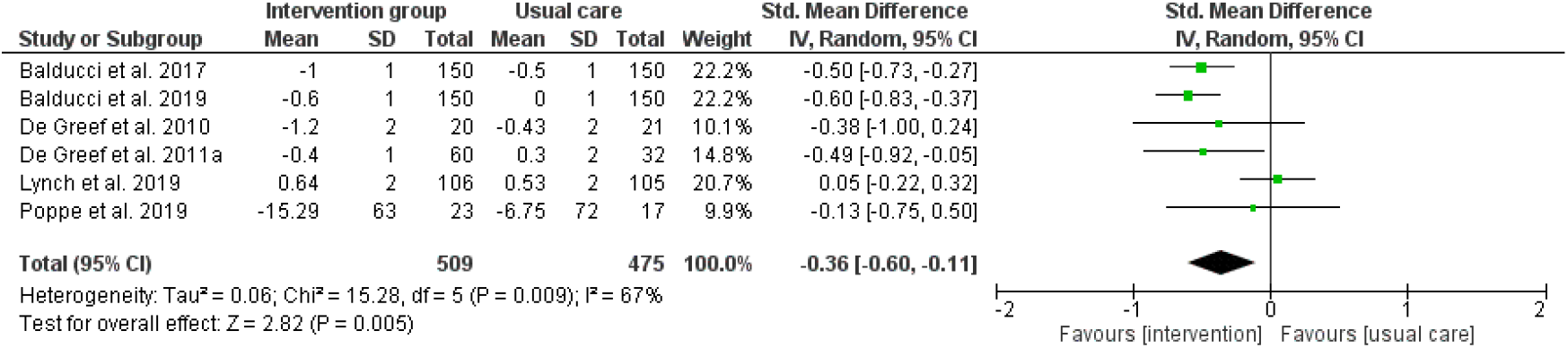
Forest plot of the effects of BCTs in the cluster ‘social support’ on sedentary time.

## Discussion

This systematic review and meta-analysis synthesized evidence from 66 randomized and non-randomized controlled trials to evaluate the effectiveness of behavior change interventions targeting PA, SB, and sleep in adults with T2D. The findings indicate that such interventions can meaningfully improve several components of 24h-MBs, with the strongest and most consistent benefits observed for PA and SB. However, it should be noted that almost no study focused on sleep which limits drawing conclusions.

Across studies, behavior change interventions produced significant improvements in multiple PA outcomes. The most consistent finding was an increase of nearly +2000 steps/day, a magnitude considered clinically relevant given evidence that adding as little as +1000 steps/day is associated with reduced mortality risk in adults with chronic diseases (86). The non-significant pooled effects for walking, despite walking being a central target of many interventions, may reflect heterogeneity in intervention designs, walking prescriptions, and measurement techniques. However, this is in contrast with the studies looking at steps/day as an outcome since many interventions integrating walking have steps/day as an outcome and not minutes or hours of walking/day. Only few studies looked at outcomes requiring sustained higher intensity (such as vigorous PA and structured exercise) and these were only significant before sensitivity analyses. This suggests that these outcomes may be more affected by adherence challenges, intervention fidelity, or study quality. In addition, there is a large heterogeneity in the measurement and conceptualization of concepts such as exercise and vigorous PA (87). Furthermore, people with T2D often have lower exercise capacity and experience more barriers than healthy people to engage in activities of higher intensity (4, 88). The mean age of participants in the current systematic review and meta-analysis is 54 years old, which is more or less the age at which a decline in higher-intensity activity is apparent in people with T2D (89). Overall, these findings support that everyday movement (e.g., steps) showing particularly robust improvements and could be an ideal candidate to focus on in behavior change interventions in the T2D population.

Some interventions also significantly reduced sedentary time. While heterogeneity was high, the direction of effect across most studies was consistent, reinforcing that SB is a modifiable behavior in this population. Study heterogeneity likely resulted from differences in SB measurement (device-based vs. self-report), intervention intensity, and frequency of prompts or feedback.

Sleep outcomes were less commonly measured, limiting conclusions. Behavior change interventions modestly improved sleep quality, but sleep duration did not significantly change. Improving sleep duration or sleep quality may require direct sleep-specific strategies such as stimulus control or sleep hygiene, rather than indirect approaches via PA or SB modification which were implemented for example in the study of Hirosaki et al. (2023), Cassidy et al. (2023), and Sinclair et al. (2023). This is consistent with literature showing that sleep improvements often require targeted behavioral strategies such as cognitive behavioral therapy for insomnia rather than broad lifestyle interventions (90). However, it should be acknowledged that only two studies were included in which sleep duration was an outcome but not the main focus of the intervention, which means that results should be interpreted with caution. The limited number of sleep-focused studies highlights an important gap, especially given the known associations between poor sleep and poorer glycemic control in T2D (91), but also the U-shaped pattern between short and long sleep duration and the association with health outcomes (6). Adults are encouraged to sleep between 7-9 hours/night, and older adults between 7-8 hours/night. This means that people with T2D having a short sleep duration (i.e., shorter than 7 hours/night) should be encouraged to sleep longer, but people with T2D with a long sleep duration (i.e., longer than 8 or 9 hours/night; depending on the age group) should be encouraged to sleep shorter. However, this is very personal and people should search for their own optimal sleep duration, which should be of high quality.

Subgroup analyses revealed that clusters of BCTs influenced intervention effectiveness. BCTs such as goal setting, action planning, and problem solving were associated with significant improvements in steps/day, total PA, MVPA, and SB. Self-monitoring and feedback - often delivered via wearables or digital tools - showed strong and consistent effects on PA and SB. This aligns with recent research demonstrating that device-based feedback produces large increases in steps/day and PA in adults with T2D (14). The systematic review of Van Rhoon et al. (2020), although looking at T2D prevention interventions, concluded that the integration of specific BCTs and digital features may optimize digital diabetes prevention interventions (92). However, effects were inconsistent for certain total PA metrics, possibly due to the reliance on different measures of PA such as device-based measurement or subjective self-reports. Social support improved steps/day, MVPA, and SB, underscoring the importance of interpersonal influences in diabetes self-management. Prior literature similarly highlights the role of social support but calls for better operationalization in intervention descriptions (93). In addition, the systematic review and network meta-analysis of Zhao et al. (2024) showed that not the number of BCTs improves effectiveness of interventions focusing on increasing PA in people with T2D but rather the use of BCTs in general and their application or fidelity when used in interventions (13). In the same systematic review, increases in PA also induced positive effects on health outcomes such as a reduction in HbA1c when goal setting was used, demonstrating that changing behavior also changes health.

The significant improvements that were found in for example MVPA, LPA or SB might be translated to studies using compositional data analysis looking into how reallocating time from one behavior to another has a theoretical improvement of cardiometabolic health outcomes. For example, theoretically reallocating time from SB to LPA or MVPA improves cardiometabolic health outcomes in T2D (4), and breaking up prolonged sitting improves postprandial glucose, insulin sensitivity, and cardiometabolic markers in T2D (94). However, these theoretical reallocations of time and their effects on health outcomes need to be confirmed in experimental studies focusing on the full 24-hour composition. Using BCTs in those interventions might increase the likelihood of being effective.

The evidence supports the use of goal setting, action planning, self-monitoring, feedback, and social support as effective BCTs to change behaviors of people with T2D into diabetes lifestyle counseling. Interventions using wearable devices or step-based feedback appear particularly feasible and effective. In the future, interventions could target 24-h MBs, recognizing the interdependence of PA, SB and sleep. Furthermore, it could be of added value to qualitatively explore how BCTs can be integrated into diabetes lifestyle counseling. In addition, more future studies should focus on the inclusion of sleep (sleep duration, sleep quality, chronotype) to address the current gap in literature.

One of the strengths of the current systematic review and meta-analysis was the use of the PRISMA guidelines to systematically report about the different steps performed. In addition, the inclusion of different behaviors tried to provide a broad overview of the current literature on the effects of interventions targeting PA, SB and sleep. However, there were also some limitations. Heterogeneity across studies was substantial, reflecting variation in intervention duration, intensity, delivery mode, and measurement tools. Risk of bias assessments indicated concerns in randomization, blinding, and selective reporting in some studies. Furthermore, long-term follow-up was limited, restricting conclusions about maintenance of behavior change which is an essential element in chronic disease management. Variability in measurement tools (accelerometry vs. self-report) likely contributed to inconsistent effect sizes for certain outcomes. Future studies using BCTs should report these more explicitly in their methodology, since in more than half of the studies the BCTs were assigned by the authors themselves which might have caused bias.

## Conclusion

This systematic review and meta-analysis demonstrates that behavior change interventions significantly improve PA, reduce SB, and enhance sleep quality while sleep duration remains insufficiently addressed in adults with T2D. BCT clusters involving goal setting, monitoring, feedback, and social support appear particularly effective. The overall findings highlight the value of theory-informed interventions for improving 24-hour movement behaviors and supporting diabetes self-management.

## Supporting information

Additional file 1: search strategy

Additional file 2: data-extraction

Additional file 3: BCTs

Additional file 4: risk of bias

## Data Availability

All data produced in the present work are contained in the manuscript

