## Additional file 1: search strategy for "The effect of lifestyle interventions using behavior change techniques to improve physical activity, sedentary behavior and/or sleep in adults with type 2 diabetes mellitus: a systematic review and meta-analysis of randomized controlled trials"

| | | |
|---|---|---|
| Pubmed | ((“Diabetes Mellitus, type 2”[Mesh] OR “Diabetes Mellitus type 2”[TIAB] OR “Diabetes type 2”[TIAB] OR “Type 2 Diabetes”[TIAB] OR “Diabetes Mellitus type II”[TIAB] OR “Diabetes type II”[TIAB] OR “type II diabetes”[TIAB] OR “Diabetes Mellitus type ii”[TIAB] OR “Diabetes type ii”[TIAB] OR “type ii diabetes”[TIAB] OR “Non-Insulin-Dependent Diabetes”[TIAB] OR “Non-Insulin Dependent Diabetes”[TIAB] OR “non insulin dependent diabetes”[TIAB] OR “Noninsulin Dependent Diabetes”[TIAB] OR “Noninsulin-Dependent Diabetes”[TIAB] OR “NIDDM”[TIAB] OR “Stable Diabetes”[TIAB] OR “adults onset diabetes”[TIAB] OR “adult-onset diabetes”[TIAB] OR “dm 2”[TIAB] OR “T2DM”[TIAB] OR “DMT2”[TIAB] OR “T2D”[TIAB] OR “DT2”[TIAB])<br>**AND**<br>(“Exercise”[Mesh] OR “Exercise”[TIAB] OR “Physical activity”[TIAB] OR “Physical activities”[TIAB] OR “walking”[Mesh] OR “walking”[TIAB] OR “walk”[TIAB] OR “Running”[Mesh] OR “Running”[TIAB] OR “Run”[TIAB] OR “Leisure activity”[TIAB] OR “Activities of Daily Living” [MeSH] OR “Leisure Activities”[Mesh] OR “Leisure Activities”[TIAB] OR “sport”[TIAB] OR “Sports”[Mesh] OR “Sports”[TIAB] OR “Motor Activity” [MeSH] OR “motor activity”[TIAB] OR “motor activities”[TIAB] OR “Locomotor activity”[TIAB] OR “locomotor activities”[TIAB] OR “Physical movement”[TIAB] OR “Physical movements”[TIAB] OR “Physical performance”[TIAB] OR “physical performances”[TIAB] OR “Physical effort”[TIAB] OR “Physical efforts”[TIAB] OR “Break”[TIAB] OR “Breaks”[TIAB] OR “Sedentary Behaviour”[TIAB] OR “Sedentary Behaviours”[TIAB] OR “Sedentary Behavior”[Mesh] OR “Sedentary Behavior”[TIAB] OR “Sedentary Behaviours”[TIAB] OR “Sedentary”[TIAB] OR “Sedentariness”[TIAB] OR “Physical inactivity”[TIAB] OR “Lack of physical activity”[TIAB] OR “sitting”[TIAB] OR “sit”[TIAB] OR “Stationary”[TIAB] OR “active transport”[TIAB] OR “passive transport”[TIAB] OR “Motor transport”[TIAB] OR “Screen Time” [MeSH] “Screen time”[TIAB] OR “Screen use”[TIAB] OR “Television”[TIAB] OR “TV”[TIAB] OR “Tablet”[TIAB] OR “Smartphone”[TIAB] OR “Reading”[TIAB] OR “sleep”[Mesh] OR “sleep”[TIAB] OR “sleeping”[TIAB] OR “lying”[TIAB] OR “Nap”[TIAB] OR “Napping”[TIAB] OR “sleepiness”[TIAB] OR “Healthy Lifestyle” [MeSH])<br>**AND**<br>(“Randomized controlled trial” [MESH] OR “randomized controlled trial” [TIAB] OR “randomised controlled trial” [TIAB] OR “RCT” [TIAB] OR “Clinical Trial” [MESH] OR “clinical trial” [TIAB] OR “clinical study” [TIAB] OR “Clinical | 1011 hits |

| | | |
|---|---|---|
| | Study” [MESH] OR “intervention” [TIAB] OR “program” [TIAB] OR “multicomponent” [TIAB] OR “behavior change” [TIAB] OR “behavioral change” [TIAB])<br>**NOT**<br>(“animal” [TIAB] OR “gestational” [TIAB] OR “mice” [TIAB]) | |
| Web of Science | TS=(("Diabetes Mellitus type 2" OR "Diabetes type 2" OR "Type 2 Diabetes" OR "Diabetes Mellitus type II" OR "Diabetes type II" OR "type II diabetes" OR "Diabetes Mellitus type ii" OR "Diabetes type ii" OR "type ii diabetes" OR "Non-Insulin-Dependent Diabetes" OR "Non-Insulin Dependent Diabetes" OR "non insulin dependent diabetes" OR "Noninsulin Dependent Diabetes" OR "Noninsulin-Dependent Diabetes" OR "NIDDM" OR "Stable Diabetes" OR "adults onset diabetes" OR "adult-onset diabetes" OR "dm 2" OR "T2DM" OR "DMT2" OR "T2D" OR "DT2") AND ("Exercise” OR "Exercise" OR "Physical activity" OR "Physical activities" OR "walking" OR "walking" OR "walk" OR "Running" OR "Running" OR "Run" OR "Leisure activity" OR "Activities of Daily Living" OR "Leisure Activities" OR "Leisure Activities" OR "sport" OR "Sports" OR "Sports" OR "Motor Activity" OR "motor activity" OR "motor activities" OR "Locomotor activity" OR "locomotor activities" OR "Physical movement" OR "Physical movements" OR "Physical performance" OR "physical performances" OR "Physical effort" OR "Physical efforts" OR "Break" OR "Breaks" OR "Sedentary Behaviour" OR "Sedentary Behaviours" OR "Sedentary Behavior" OR "Sedentary Behavior" OR "Sedentary Behaviours" OR "Sedentary" OR "Sedentariness" OR "Physical inactivity" OR "Lack of physical activity" OR "sitting" OR "sit" OR "Stationary" OR "active transport" OR "passive transport" OR "Motor transport" OR "Screen Time" "Screen time" OR "Screen use" OR "Television" OR "TV" OR "Tablet" OR "Smartphone" OR "Reading" OR "sleep" OR "sleep" OR "sleeping" OR "lying" OR "Nap" OR "Napping" OR "sleepiness" OR "Healthy Lifestyle") AND ("Randomized controlled trial" OR "randomized controlled trial" OR "randomised controlled trial" OR "RCT" OR "Clinical Trial" OR "clinical trial" OR "clinical study" OR "Clinical Study" OR "intervention" OR "program" OR "multicomponent" OR "behavior change" OR "behavioral change" ) NOT (“animal” OR “gestational” OR “mice”)) | 7275 hits |
| Embase | ((‘Diabetes Mellitus type 2’:ti,ab,kw OR ‘Diabetes type 2’:ti,ab,kw OR ‘Type 2 Diabetes’:ti,ab,kw OR ‘Diabetes Mellitus type II’:ti,ab,kw OR ‘Diabetes type II’:ti,ab,kw OR ‘type II diabetes’:ti,ab,kw OR ‘Diabetes Mellitus type ii’:ti,ab,kw OR ‘Diabetes type ii’:ti,ab,kw OR ‘type ii diabetes’:ti,ab,kw OR ‘Non-Insulin-Dependent Diabetes’:ti,ab,kw OR ‘Non-Insulin Dependent Diabetes’:ti,ab,kw OR ‘non insulin dependent diabetes’:ti,ab,kw OR ‘Noninsulin Dependent Diabetes’:ti,ab,kw OR ‘Noninsulin-Dependent Diabetes’:ti,ab,kw OR ‘NIDDM’:ti,ab,kw OR ‘Stable Diabetes’:ti,ab,kw OR ‘adults onset diabetes’:ti,ab,kw OR ‘adult-onset diabetes’:ti,ab,kw OR ‘dm 2’:ti,ab,kw OR ‘T2DM’:ti,ab,kw OR ‘DMT2’:ti,ab,kw OR ‘T2D’:ti,ab,kw OR ‘DT2’:ti,ab,kw) AND (‘Exercise’:ti,ab,kw OR ‘Exercise’:ti,ab,kw OR ‘Physical activity’:ti,ab,kw OR ‘Physical activities’:ti,ab,kw OR ‘walking’:ti,ab,kw OR | 8709 hits |

| | | |
|---|---|---|
| | ‘walking’:ti,ab,kw OR ‘walk’:ti,ab,kw OR ‘Running’:ti,ab,kw OR ‘Running’:ti,ab,kw OR ‘Run’:ti,ab,kw OR ‘Leisure activity’:ti,ab,kw OR ‘Activities of Daily Living’:ti,ab,kw OR ‘Leisure Activities’:ti,ab,kw OR ‘Leisure Activities’:ti,ab,kw OR ‘sport’:ti,ab,kw OR ‘Sports’:ti,ab,kw OR ‘Sports’:ti,ab,kw OR ‘Motor Activity’:ti,ab,kw OR ‘motor activity’:ti,ab,kw OR ‘motor activities’:ti,ab,kw OR ‘Locomotor activity’:ti,ab,kw OR ‘locomotor activities’:ti,ab,kw OR ‘Physical movement’:ti,ab,kw OR ‘Physical movements’:ti,ab,kw OR ‘Physical performance’:ti,ab,kw OR ‘physical performances’:ti,ab,kw OR ‘Physical effort’:ti,ab,kw OR ‘Physical efforts’:ti,ab,kw OR ‘Break’:ti,ab,kw OR ‘Breaks’:ti,ab,kw OR ‘Sedentary Behaviour’:ti,ab,kw OR ‘Sedentary Behaviours’:ti,ab,kw OR ‘Sedentary Behavior’:ti,ab,kw OR ‘Sedentary Behavior’:ti,ab,kw OR ‘Sedentary Behaviours’:ti,ab,kw OR ‘Sedentary’:ti,ab,kw OR ‘Sedentariness’:ti,ab,kw OR ‘Physical inactivity’:ti,ab,kw OR ‘Lack of physical activity’:ti,ab,kw OR ‘sitting’:ti,ab,kw OR ‘sit’:ti,ab,kw OR ‘Stationary’:ti,ab,kw OR ‘active transport’:ti,ab,kw OR ‘passive transport’:ti,ab,kw OR ‘Motor transport’:ti,ab,kw OR ‘Screen Time’:ti,ab,kw OR ‘Screen use’:ti,ab,kw OR ‘Television’:ti,ab,kw OR ‘TV’:ti,ab,kw OR ‘Tablet’:ti,ab,kw OR ‘Smartphone’:ti,ab,kw OR ‘Reading’:ti,ab,kw OR ‘sleep’:ti,ab,kw OR ‘sleeping’:ti,ab,kw OR ‘lying’:ti,ab,kw OR ‘Nap’:ti,ab,kw OR ‘Napping’:ti,ab,kw OR ‘sleepiness’:ti,ab,kw OR ‘Healthy Lifestyle’:ti,ab,kw) AND (‘Randomized controlled trial’:ti,ab,kw OR ‘randomised controlled trial’:ti,ab,kw OR ‘RCT’:ti,ab,kw OR ‘Clinical Trial’:ti,ab,kw OR ‘clinical trial’:ti,ab,kw OR ‘clinical study’:ti,ab,kw OR ‘Clinical Study’:ti,ab,kw OR ‘intervention’:ti,ab,kw OR ‘program’:ti,ab,kw OR ‘multicomponent’:ti,ab,kw OR ‘behavior change’:ti,ab,kw OR ‘behavioral change’:ti,ab,kw ) NOT (‘animal’:ti,ab,kw OR ‘gestational’:ti,ab,kw OR ‘mice’:ti,ab,kw)) | |
| Scopus | Optie 1<br><br>TITLE-ABS-KEY(("Diabetes Mellitus type 2" OR "Diabetes type 2" OR "Type 2 Diabetes" OR "Diabetes Mellitus type II" OR "Diabetes type II" OR "type II diabetes" OR "Diabetes Mellitus type ii" OR "Diabetes type ii" OR "type ii diabetes" OR "Non-Insulin-Dependent Diabetes" OR "Non-Insulin Dependent Diabetes" OR "non insulin dependent diabetes" OR "Noninsulin Dependent Diabetes" OR "Noninsulin-Dependent Diabetes" OR "NIDDM" OR "Stable Diabetes" OR "adults onset diabetes" OR "adult-onset diabetes" OR "dm 2" OR "T2DM" OR "DMT2" OR "T2D" OR "DT2") AND ("Exercise” OR "Exercise" OR "Physical activity" OR "Physical activities" OR "walking" OR "walking" OR "walk" OR "Running" OR "Running" OR "Run" OR "Leisure activity" OR "Activities of Daily Living" OR "Leisure Activities" OR "Leisure Activities" OR "sport" OR "Sports" OR "Sports" OR "Motor Activity" OR "motor activity" OR "motor activities" OR "Locomotor activity" OR "locomotor activities" OR "Physical movement" OR "Physical movements" OR "Physical performance" OR "physical performances" OR "Physical effort" OR "Physical efforts" OR "Break" OR "Breaks" OR "Sedentary Behaviour" OR "Sedentary Behaviours" OR "Sedentary Behavior" OR "Sedentary Behavior" | 204 hits |

| | | |
|---|---|---|
| | OR "Sedentary Behaviours" OR "Sedentary" OR "Sedentariness" OR "Physical inactivity" OR "Lack of physical activity" OR "sitting" OR "sit" OR "Stationary" OR "active transport" OR "passive transport" OR "Motor transport" OR "Screen Time" "Screen time" OR "Screen use" OR "Television" OR "TV" OR "Tablet" OR "Smartphone" OR "Reading" OR "sleep" OR "sleep" OR "sleeping" OR "lying" OR "Nap" OR "Napping" OR "sleepiness" OR "Healthy Lifestyle") AND ("Randomized controlled trial" OR "randomized controlled trial" OR "randomised controlled trial" OR "RCT" OR "Clinical Trial" OR "clinical trial" OR "clinical study" OR "Clinical Study" OR "intervention" OR "program" ) AND ("multicomponent" OR "behavior change" OR "behavioral change" ))<br><br>Optie 2<br><br>TITLE-ABS-KEY(("Diabetes Mellitus type 2" OR "Diabetes type 2" OR "Type 2 Diabetes" OR "Diabetes Mellitus type II" OR "Diabetes type II" OR "type II diabetes" OR "Diabetes Mellitus type ii" OR "Diabetes type ii" OR "type ii diabetes" OR "Non-Insulin-Dependent Diabetes" OR "Non-Insulin Dependent Diabetes" OR "non insulin dependent diabetes" OR "Noninsulin Dependent Diabetes" OR "Noninsulin-Dependent Diabetes" OR "NIDDM" OR "Stable Diabetes" OR "adults onset diabetes" OR "adult-onset diabetes" OR "dm 2" OR "T2DM" OR "DMT2" OR "T2D" OR "DT2") AND ("Exercise” OR "Exercise" OR "Physical activity" OR "Physical activities" OR "walking" OR "walking" OR "walk" OR "Running" OR "Running" OR "Run" OR "Leisure activity" OR "Activities of Daily Living" OR "Leisure Activities" OR "Leisure Activities" OR "sport" OR "Sports" OR "Sports" OR "Motor Activity" OR "motor activity" OR "motor activities" OR "Locomotor activity" OR "locomotor activities" OR "Physical movement" OR "Physical movements" OR "Physical performance" OR "physical performances" OR "Physical effort" OR "Physical efforts" OR "Break" OR "Breaks" OR "Sedentary Behaviour" OR "Sedentary Behaviours" OR "Sedentary Behavior" OR "Sedentary Behavior" OR "Sedentary Behaviours" OR "Sedentary" OR "Sedentariness" OR "Physical inactivity" OR "Lack of physical activity" OR "sitting" OR "sit" OR "Stationary" OR "active transport" OR "passive transport" OR "Motor transport" OR "Screen Time" "Screen time" OR "Screen use" OR "Television" OR "TV" OR "Tablet" OR "Smartphone" OR "Reading" OR "sleep" OR "sleep" OR "sleeping" OR "lying" OR "Nap" OR "Napping" OR "sleepiness" OR "Healthy Lifestyle") AND ("Randomized controlled trial" OR "randomized controlled trial" OR "randomised controlled trial" OR "RCT" OR "Clinical Trial" OR "clinical trial" OR "clinical study" OR "Clinical Study" OR "intervention" OR "program" OR "multicomponent" OR "behavior change" OR "behavioral change") AND NOT (“animal” OR “gestational” OR “mice”)) | 2116 hits 14/12/2023 |
