## Additional file 4: risk of bias for "The effect of lifestyle interventions using behavior change techniques to improve physical activity, sedentary behavior and/or sleep in adults with type 2 diabetes mellitus: a systematic review and meta-analysis of randomized controlled trials"

### Supplementary file 3 – Risk of bias assessment

**Revised Cochrane risk-of-bias tool for randomized trials**

<u>Domains</u>

| | |
|---|---|
| Domain 1 | Randomisation process |
| Domain 2 | Deviations from the intended interventions |
| Domain 3 | Missing outcome data |
| Domain 4 | Measurement of the outcome |
| Domain 5 | Selection of reported results |

<u>Legend</u>

| | |
|---|---|
| + | Low risk of bias |
| ! | Some concerns |
| - | High risk of bias |

| **Author et al. year** | **Domain 1** | **Domain 2** | **Domain 3** | **Domain 4** | **Domain 5** | **Overall bias** |
|---|---|---|---|---|---|---|
| Alghafri *et al.* 2018 | - | - | - | ! | ! | - |
| Alonso-DomÍnguez *et al.* 2019 | + | + | + | + | + | + |
| Araiza *et al.* 2006 | ! | + | ! | + | + | ! |
| Balducci *et al.* 2019 | + | + | + | + | + | + |
| Balducci *et al.* 2017 | + | + | + | + | + | + |
| Beverly *et al.* 2013 | + | + | + | + | + | + |
| Bjorgaas et al. 2005 | + | ! | + | + | + | ! |
| Cassidy *et al.* 2023 | ! | ! | + | + | + | ! |
| Christian *et al.* 2008 | + | ! | + | ! | + | ! |

| | | | | | | |
|---|---|---|---|---|---|---|
| Akinci *et al.* 2018 | + | + | ! | + | + | ! |
| Andrews *et al.* 2011 | + | + | + | + | + | + |
| De Greef *et al.* 2011a | + | + | + | + | + | + |
| De Greef *et al.* 2010 | + | + | + | + | + | + |
| De Greef *et al.* 2011b | + | + | + | + | + | + |
| Dyson *et al.* 2010 | + | + | + | + | ! | ! |
| Eakin *et al.* 2013 | + | + | + | + | + | + |
| Fayehun *et al.* 2018 | + | + | + | + | + | + |
| Fritz *et al.* 2006 | - | + | + | - | ! | - |
| Gray *et al.* 2021 | + | + | + | + | + | + |
| Hashim *et al.* 2021 | ! | + | + | + | + | ! |
| Poppe *et al.* 2019 | + | + | + | + | + | + |
| Samuel-Hodge *et al.* 2017 | + | - | + | ! | + | - |
| Serin & Saritas, 2020 | + | + | + | ! | + | ! |
| Sinclair *et al.* 2023 | + | + | + | + | + | + |
| Sung & Bae, 2012 | + | - | + | + | ! | - |
| Thuita *et al.* 2020 | + | + | + | ! | ! | ! |
| Toobert *et al.* 2007 | + | + | + | ! | + | + |
| Tudor-Locke *et al.* 2004 | + | + | + | + | ! | ! |

| | | | | | | |
|---|---|---|---|---|---|---|
| Unick *et al.* 2016 | + | - | + | + | ! | - |
| Van Dyck *et al.* 2013 | + | + | + | ! | + | ! |
| Varney *et al.* 2014 | ! | + | + | ! | + | ! |
| Vluggen *et al.* 2021 | + | - | + | ! | + | - |
| Welschen *et al.* 2013 | + | + | + | ! | + | + |
| Yoo *et al.* 2008 | + | + | + | ! | + | ! |
| Zhang *et al.* 2021 | + | + | + | ! | + | ! |
| Zuo *et al.* 2020 | + | + | + | ! | + | ! |
| Glasgow *et al.* 2010 | + | ! | + | - | + | - |
| Lorig *et al*. 2009 | ! | - | + | - | + | - |
| Hirosaki *et al.* 2023 | + | + | + | ! | + | ! |
| Hordern *et al.* 2009 | + | + | ! | ! | + | ! |
| Irwig *et al*. 2012 | + | + | + | ! | + | ! |
| Jayasree & Stalin, 2019 | + | + | + | ! | ! | ! |
| Kattelmann *et al.* 2009 | + | + | - | ! | + | - |
| Keyserling et al. 2002 | + | ! | + | + | + | ! |
| Khosravan *et al.* 2015 | + | + | + | ! | + | ! |
| Kim & Kang, 2006 | + | + | + | ! | + | ! |
| King *et al.* 2006 | + | + | + | ! | + | ! |

| | | | | | | |
|---|---|---|---|---|---|---|
| Kirk *et al.* 2009 | + | + | + | + | + | + |
| Jakicic *et al.* 2009 | + | + | + | ! | + | ! |
| Lari et al. 2018 | + | + | + | + | + | + |
| Lehmann *et al.* 1995 | + | - | + | + | + | ! |
| Li *et al.* 2023 | + | + | + | + | + | + |
| Liebrich *et al.* 2009 | + | + | ! | + | + | ! |
| Liu *et al.* 2012 | + | + | ! | + | + | ! |
| Lorig *et al.* 2010 | + | ! | ! | + | + | ! |
| Ma *et al.* 2008 | + | ! | + | + | + | ! |
| McEwen *et al.* 2017 | + | ! | - | + | + | ! |
| Miyamoto *et al.* 2017 | + | + | + | + | + | + |
| Morowatisharifabad *et al.* 2021 | + | + | + | + | + | + |
| Negri *et al.* 2010 | + | + | + | + | + | + |
| Nepper *et al.* 2019 | - | + | + | + | + | - |
| Piette *et al.* 2011 | + | + | + | + | + | + |
| Plotnikoff *et al.* 2013 | + | + | + | + | + | + |
| Mahdizadeh *et al.* 2013 | - | + | + | + | - | - |
| Lee *et al.* 2015 | + | ! | + | ! | + | ! |
| Lynch *et al.* 2019 | + | + | + | + | + | + |

| | | | | | | |
|---|---|---|---|---|---|---|
| Matsushita *et al.* 2022 | + | + | + | + | + | + |
| Sazlina *et al.* 2015 | + | - | + | + | + | ! |

**Total**

| | |
|---|---|
| + | **27** |
| ! | **38** |
| - | **11** |
